# Comparative Effectiveness of Single vs. Dual WhatsApp Reminders on No-shows: A Target Trial Emulation within the Public Health System of Buenos Aires, Argentina

**DOI:** 10.64898/2026.08.17.26360609

**Authors:** Santiago Esteban, Gastón Quintana, Mario Sanchez, Adolfo Rubinstein, Alejandro Szmulewicz

**Affiliations:** Centro de Implementación e Innovación en Políticas de Salud (CIIPS), Instituto de Efectividad Clínica y Sanitaria (IECS), Buenos Aires, Argentina; Gerencia Operativa de Gestión de Información y Estadísticas de Salud, Ministerio de Salud de la Ciudad de Buenos Aires, Buenos Aires, Argentina; Banco Interamericano de Desarrollo, Buenos Aires, Argentina; Department of Epidemiology, Harvard TH Chan School of Public Health, Boston, MA 02115, USA

**Keywords:** No-shows, absenteeism, non-attendance, reminders, outpatient, WhatsApp, Target trial emulation

## Abstract

**Background:** Digital reminders reduce outpatient no-shows, but the optimal timing and frequency of messages remain unclear, particularly in Latin American public health systems. We emulated a target trial to evaluate the comparative effectiveness of four WhatsApp reminder strategies on appointment absenteeism and patient-initiated cancellations.

**Methods:** We analyzed administrative and electronic health-record data from the public health system of the Autonomous City of Buenos Aires, Argentina (June 2023–May 2024). Eligible individuals had scheduled an in-person outpatient appointment in one of 15 prioritized specialties at least 75 hours in advance and had a mobile phone on record. We compared four strategies: (1) dual reminders at ∼72 and ∼24 hours before the appointment; (2) a single reminder at ∼72 hours; (3) a single reminder at ∼24 hours; and (4) no reminders. The primary outcome was the proportion of no-shows by the end of follow-up. Secondary outcomes were the cumulative incidence of patient-initiated cancellations overall, within 12 hours of the appointment, and followed by rebooking. We emulated the target trial using a cloning-censoring-weighting approach to estimate the per-protocol effect, with inverse-probability weights to address time-varying confounding and selection bias. Cumulative incidence of secondary outcomes was estimated using weighted Kaplan-Meier curves. Three pre-specified sensitivity analyses and standardized mean differences assessed robustness and covariate balance.

**Results:** A total of 475,214 first eligible person-appointments were included; baseline no-show risk in the control arm was 34.6%. All three active strategies reduced no-shows compared with no reminders. The single 24-hour reminder produced the largest reduction (Risk Ratio [RR] 0.76, 95% CI 0.72–0.81; Risk Difference [RD] −8.21 percentage points [pp], 95% CI −9.68, −6.54), followed by the dual-reminder strategy (RR 0.80, 95% CI 0.79-0.81; RD −7.05 pp, 95% CI −7.41, −6.71) and the single 72-hour reminder (RR 0.91, 95% CI 0.84–0.99; RD −3.16 pp, 95% CI −5.69, −0.49). All active strategies increased patient-initiated cancellations relative to control, with the dual-reminder strategy producing the largest increase. Sensitivity analyses preserved the qualitative ranking of strategies across all specifications.

**Conclusions:** In this large target trial emulation, a single just-in-time WhatsApp reminder sent ∼24 hours before the appointment was as effective as a dual-reminder schedule in preventing no-shows and superior to a distal 72-hour reminder alone. Adding a second, distal reminder provided no measurable benefit for attendance but substantially increased patient-initiated cancellations, which may be operationally valuable when active slot reallocation is a goal. These findings support timing, rather than frequency, as the primary lever of digital-reminder effectiveness, and favor the deployment of a single proximal reminder as the default strategy in resource-constrained outpatient settings.

## Background

Medical appointment no-shows, missing a scheduled consultation without prior cancellation, are a widespread challenge, with global outpatient rates ranging from 15% to 30% [1–5]. These missed appointments severely disrupt healthcare efficiency by increasing operational costs, wasting clinical resources, and reducing overall access to care [6,7]. Furthermore, no-shows compromise patient outcomes by delaying chronic disease management, which can lead to preventable clinical complications, increased morbidity, and costly hospitalizations [6–8].

Forgetting an appointment is frequently cited as a primary cause of absenteeism [9,10]. From an epidemiological and behavioral perspective, a no-show is not merely a random event, but rather a breakdown in the continuum of care that can be analyzed through the Transtheoretical Model of Change [11]. In the healthcare context, a patient’s intention to attend consultation must be sustained against competing daily stressors. Instant messaging and other digital notifications can be conceptualized as behavioral interventions that facilitate the transition from the preparation phase to action. Recently, instant messaging platforms like WhatsApp have gained prominence in facilitating interactions between healthcare systems and patients, including the automated delivery of reminders [12–21].

Data from randomized clinical trials show that sending multiple reminders (e.g., at 7, 3 and 1 day prior to the appointment) is significantly more effective at reducing absenteeism compared to no reminders, without negatively impacting patient satisfaction [22–24]. However, although previous trials have explored different combinations of reminder number and timing, none has compared head-to-head the four strategies of clinical interest: two reminders (3 days and 1 day prior to the appointment), a single reminder 3 days prior, a single reminder 1 day prior, and no reminder at all. As a result, the optimal frequency and timing of digital reminders remains unknown. Moreover, to the best of our knowledge, no study has evaluated WhatsApp-based reminder strategies in the Latin American public health sector. Because it is impractical and unlikely that we will have other randomized trials exploring different frequencies and timing anytime soon, we emulated a target trial comparing the effect of different WhatsApp reminder strategies on appointment absenteeism and cancellations using data from the public health system of Autonomous City of Buenos Aires (ACBA), Argentina (June 2023–May 2024).

By identifying the optimal frequency and timing for digital reminders, this study provides actionable evidence to maximize resource efficiency and long-term sustainability in a South American public healthcare setting.

## Methodology

We designed an observational study to emulate a pragmatic randomized clinical trial. For this, we first specify the pragmatic, randomized trial that would answer the causal question. Then we specify how it can be emulated with the available observational data. Table 1 summarizes the specifications of the target trial and its corresponding observational emulation.

**Table 1.** Target trial specification and emulation using observational data from the Buenos Aires City public health system databases (June 2023-May 2024)

|  |  |  |
| --- | --- | --- |
| Component | Target trial | Emulation with observational data |
| Eligibility criteria | Individuals who scheduled an in-person outpatient appointment for the period June 2023-May 2024, where:<br>□ It is an appointment for one of the prioritized specialties (cardiology, internal medicine, dermatology, endocrinology, gynecology, infectology, neurology, obstetrics, ophthalmology, oncology, otolaryngology, psychology, traumatology, family medicine/general medicine, pediatrics)<br>□ The appointment was booked at least 75 hours prior to the appointment date<br>□ The appointment was booked by a usual-user of the Buenos Aires public health system<br>□ The appointment was not cancelled prior to time zero<br>□ The user has a valid mobile phone<br>□ The user has not received a WhatsApp reminder before time zero<br>□ The appointment was booked at a hospital or primary healthcare facility | The same as the target trial, considering the following:<br>□ To determine whether an individual had a mobile phone on record, we analyzed the structure of the registered phone numbers, as mobile numbers in Argentina follow a specific format that distinguishes them from landlines.<br>□ The identification of usual system users was emulated by having at least two non-emergency consultations registered in the system in the last 2 years |
| Treatment strategies | └ Strategy 1: Two reminders, ~72 (75-64h) & ~24h (27-15h) prior to the appointment<br>└ Strategy 2: One reminder, ~72 (75-64h) prior to the appointment<br>└ Strategy 3: One reminder, ~24h (27-15h) prior to the appointment<br>└ Strategy 4: No reminders<br>Under all strategies, patients do not receive reminders outside of the specified windows and receive only one reminder in the specified window. | We utilized a database linking the dispatched WhatsApp messages to each scheduled appointment, in which the time and date of each dispatched message was recorded. Figure 1 shows the distribution of hours prior to the appointment at which a reminder was sent. No participant received more than two reminders, nor received any reminders in between reminder windows (-63, -28h). |
| Assignment procedures | 1:1:1:1 Randomization, Open label | Randomization will be emulated by cloning each individual and assigning each clone to one treatment strategy. |
| **Follow-up** | For each eligible participant, follow-up begins 75 hours before the scheduled appointment time and ends at the earliest of the following events: i) appointment, ii) loss to follow-up (defined as a healthcare system-initiated cancellation), or ii) the administrative end of the study (May 31st 2024). | Same as the target trial |
| **Results** | Primary outcome: percentage of scheduled appointments resulting in a no-show by the end of follow-up<br>Secondary outcomes:<br>Cumulative incidence of appointments canceled by the patient that are subsequently rebooked<br>Cumulative incidence of appointments canceled by the patient at 12 hours prior to the scheduled time. | Data on scheduled appointment are obtained from scheduling system that logs booked appointments, specific dates, and appointment statuses (scheduled, canceled, no-show, or attended) |
| **Causal contrasts** | Intention-to-treat (ITT) and per-protocol effect (PP) | Per-protocol effect |
| **Identifying assumptions** | In terms of loss to follow-up, we assume no loss to follow-up given the short follow-up period. Also, given the type of intervention, if there was loss to follow-up, we don't expect it to be related to the intervention.<br>For competing events (healthcare system-initiated cancellations) we assume no residual bias after adjusting for the proposed covariates using IPCW.<br>For the ITT effect: under an adequate randomization scenario, no additional assumptions are required.<br>For the PP effect: Since participants are censored when they deviate from their assigned strategy, we assume no residual bias after adjusting for the proposed covariates using IPTW. | Same as the target trial in terms of loss to follow-up, handling competing events and estimating the PP effect. |
| **Data analysis plan** | For the primary outcome, we compare the proportion of no-shows in each group by the end of the follow-up period. For secondary outcomes, non-parametric Kaplan-Meier cumulative incidence curves to estimate Risk differences and ratios<br>Subgroup analysis<br>Age group (0-12, 13-18, 19-40, 41-65 and >65 years old)<br>Booking lead time (3–15, 16–30, 31–45, and >45 days)<br>Medical specialty (Cardiology, Internal Medicine, Dermatology, Endocrinology, Gynecology, Infectious Diseases, Neurology, Obstetrics, Ophthalmology, Oncology, Otolaryngology, Psychology, Orthopedics and Traumatology, Family Medicine/General Practice, and Pediatrics.)<br>Healthcare facility type (Hospital vs Primary Care Center) | PP analysis: Same as the target trial, only that the analysis was conducted in an expanded dataset, which included clones of each individual for each strategy (e.g., each participant had four copies, one for each strategy)[31].<br>Subgroup analysis: same as the target trial. |

### Target trial specification

The target trial includes individuals of any age group who have an active schedule for an in-person outpatient appointment in the next 75 hours scheduled at a hospital or primary care health facility within the Buenos Aires City public health system for one of the following eligible medical specialties: Cardiology, Internal Medicine, Dermatology, Endocrinology, Gynecology, Infectious Diseases, Neurology, Obstetrics, Ophthalmology, Oncology, Otolaryngology, Psychology, Orthopedics and Traumatology, Family Medicine/General Practice, or Pediatrics; whose appointment was booked at least 75 hours prior to the appointment date; who were usual users of the Buenos Aires City public health system; had no prior WhatsApp reminders; and who owned a mobile phone registered in the system.

Four intervention strategies are considered: a) Strategy 1: two messages, sent approximately 72 hours (time window: 75–64 hours) and 24 hours (time window: 27–15 hours) prior to the appointment; b) Strategy 2: a single message sent 72 hours prior (time window: 75–64 hours); c) Strategy 3: a single message sent 24 hours prior (time window: 27–15 hours); and d) Strategy 4: no messages. Patients are randomly assigned to one of these four strategies at time zero and are aware of the intervention they have been assigned to.

The primary outcome is the percentage of scheduled appointments resulting in a no-show by the end of follow-up. Secondary outcomes include the cumulative incidence of: i) appointments canceled by the patient that are subsequently rebooked, and ii) overall patient-initiated cancellations and at 12 hours prior to the scheduled time (when it is realistically possible to reuse the booked time).

For each eligible participant, follow-up begins at baseline and ends at the earliest of the following events: i) appointment, ii) loss to follow-up (defined as a healthcare system-initiated cancellation), or ii) the administrative end of the study (May 31^st^ 2024).

The causal contrasts of interest are the intention-to-treat effect of being assigned to the treatment strategies and the per-protocol effect of adhering to them. Because some individuals are expected to cancel their appointment in advance during the follow-up, alternative ways to incorporate user-initiated cancellations (competing events) into the causal question may result in different numerical estimates with different interpretations. Here, we will define both causal estimands as contrasts of the marginal risk of no-shows[25].

### Data analysis plan for the target trial

The ITT analysis for the primary outcome compares the proportion of no-shows in each group by the end of follow-up. For secondary time-to-event outcomes, the analysis can be conducted non-parametrically using the Kaplan-Meier estimator and parametrically using a pooled logistic regression model that includes an indicator variable for the intervention strategy, follow-up time, and an interaction term between the two [26].

In the per-protocol analysis, individuals are censored if and when they deviate from their assigned strategy. To adjust for potential selection bias induced by this censoring, we apply time-varying inverse probability (IP) weights to each participant at each hour of follow-up, conditional on baseline covariates (sex [male, female], number of days between booking and appointment, number of appointments in the previous year, number of no-shows in the previous year, number of present appointments in the previous year, health insurance [yes, no], type of neighborhood [ACBA formal, ACBA informal, outside of ACBA, no data on neighborhood], pregnant at [yes, no], type of health facility [primary care center, specialized ambulatory center, hospital], appointment’s medical specialty, date of time zero, age in years, number of visits in the previous year, seniority within the health system in years). Tables provided in the Supplementary Material detail the censoring criteria for protocol deviations in each arm, the statistical models, and the criteria for calculating the adherence probability for each strategy. Weights are truncated at the 99th percentile to prevent extreme outliers from disproportionately influencing the results. We calculate 95% confidence intervals (CIs) using a non-parametric percentile bootstrap with 500 resamples.

Finally, we predefine four subgroup analyses: by age group (0–12, 13–18, 19–40, 41–65, and >65 years), booking lead time (3–15, 16–30, 31–45, and >45 days), type of healthcare facility (hospital vs. primary care center), and medical specialty.

### Target Trial Emulation using observational data

We emulated the abovementioned target trial using data from the transactional information systems of the public provider network of the ACBA[27]. These systems, encompassing both primary care centers and hospitals, include the appointment scheduling system, the administrative system, and the electronic health record (EHR). Demographic, insurance, contact, and geographic data are recorded at the individual patient level within the administrative system. The EHR stores clinical information, including health problems, diagnoses, and medication prescriptions. Meanwhile, the scheduling system logs booked appointments, specific dates, and appointment statuses (scheduled, canceled, no-show, or attended). Information from these three systems is integrated and linked using unique patient identifiers, consolidating daily into a centralized data warehouse. From this repository, data were extracted for the period between June 2023 and May 2024.

We used the observational data to emulate each component of the target trial as closely as possible (Table 1). Although individuals could potentially meet the eligibility criteria at several booked appointments over follow-up, we only selected the first eligible time for each individual as time zero, for computational reasons. Intervention strategies were the same as in the target trial. Because at baseline individuals had information compatible with more than one treatment strategy, we created four clones of each individual and assigned each clone to one treatment strategy [28]. This approach precludes the estimation of the intention-to-treat analysis. The per-protocol analysis proceeded exactly as described for the target trial except that it was conducted in a database with clones. All other components of the target trial were closely emulated.

### Sensitivity analysis

To assess the robustness of our findings to key analytical decisions, we performed three pre-specified sensitivity analyses. First, we varied the cut-off point used to assign each participant to a reminder strategy, examining the impact of widening or narrowing the time window around the 72-hour and 24-hour pre-appointment marks, in order to evaluate whether minor differences in the timing of message delivery relative to the protocol-defined schedule could meaningfully alter the estimated effects. Second, we re-estimated the cumulative incidence functions for the time-to-event outcomes using a parametric pooled logistic regression model weighted by the inverse-probability weights derived from the main analysis, and compared the resulting curves to those obtained from our primary non-parametric Kaplan-Meier approach; agreement between the two approaches would suggest that the parametric assumptions hold and thus the parametric estimator could be used as the main estimator for statistical efficiency. Third, we re-fit the IP weighting models using more flexible specification of the continuous variables as well as interaction terms between key baseline covariates (e.g., age, sex, and appointment specialty), to assess if model misspecification had an effect on the observed effect estimates.

For all sensitivity analyses, results were considered consistent with the primary analysis if point estimates remained within the 95% confidence intervals of the main estimates and if the qualitative conclusions regarding the relative ranking of the reminder strategies were preserved.

## Results

Figure 1 shows a flowchart of participant selection and Table 2 shows the baseline characteristics of the eligible individuals included in the emulation. Eligible participants were predominantly female (64%), with the largest age groups consisting of adults aged 41 to 65 years (30%) and young adults aged 19 to 40 years (27%). At 14 hours before the appointment (when all clones were unequivocally following one strategy), characteristics between groups were balanced, except for a higher proportion of patients residing outside of ACBA, more heavily concentrated in hospital settings, and a higher number of previous appointments in the no message group.

**Figure 1.**
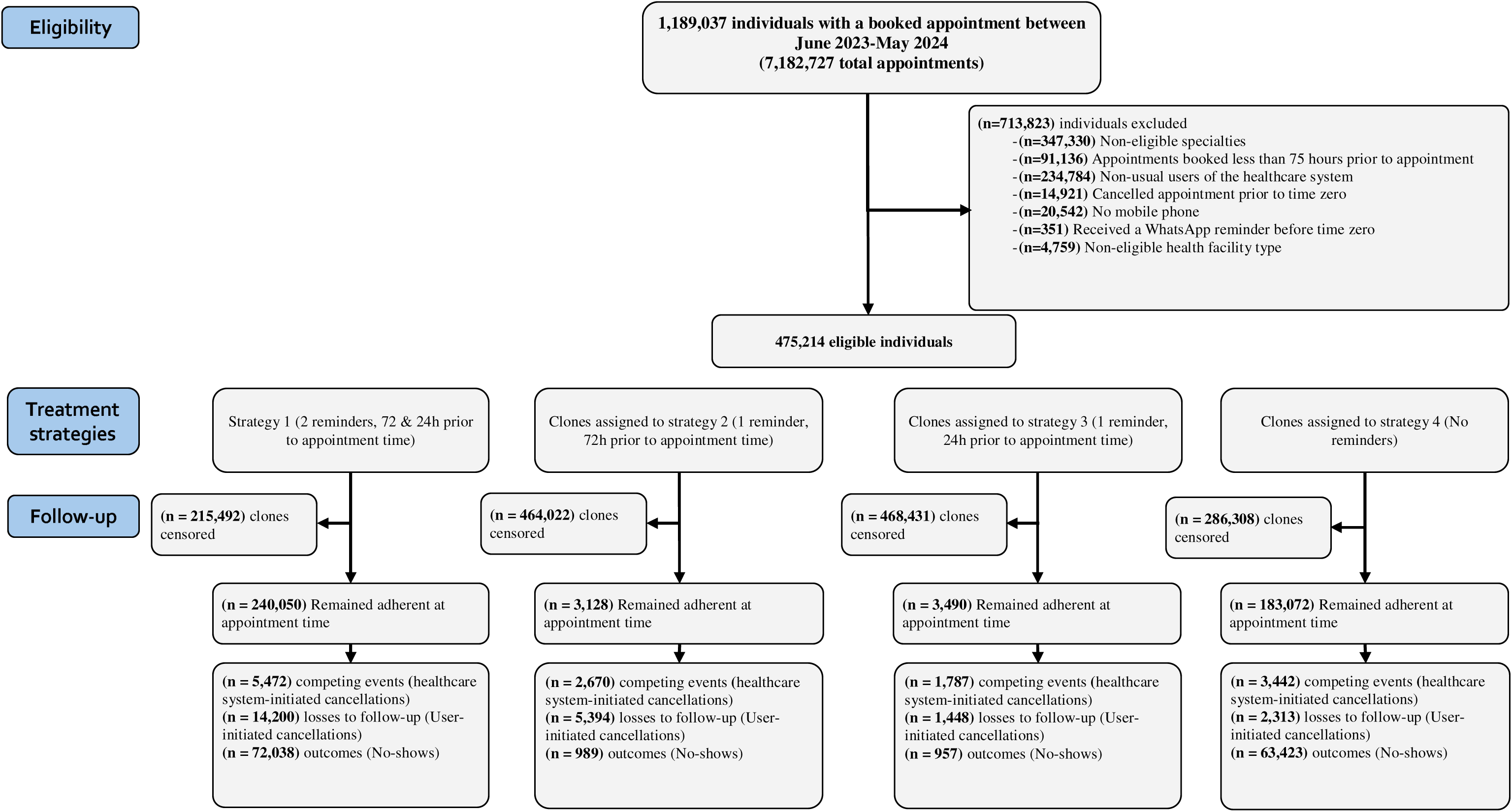
Flowchart of eligible participants.

**Table 2.**
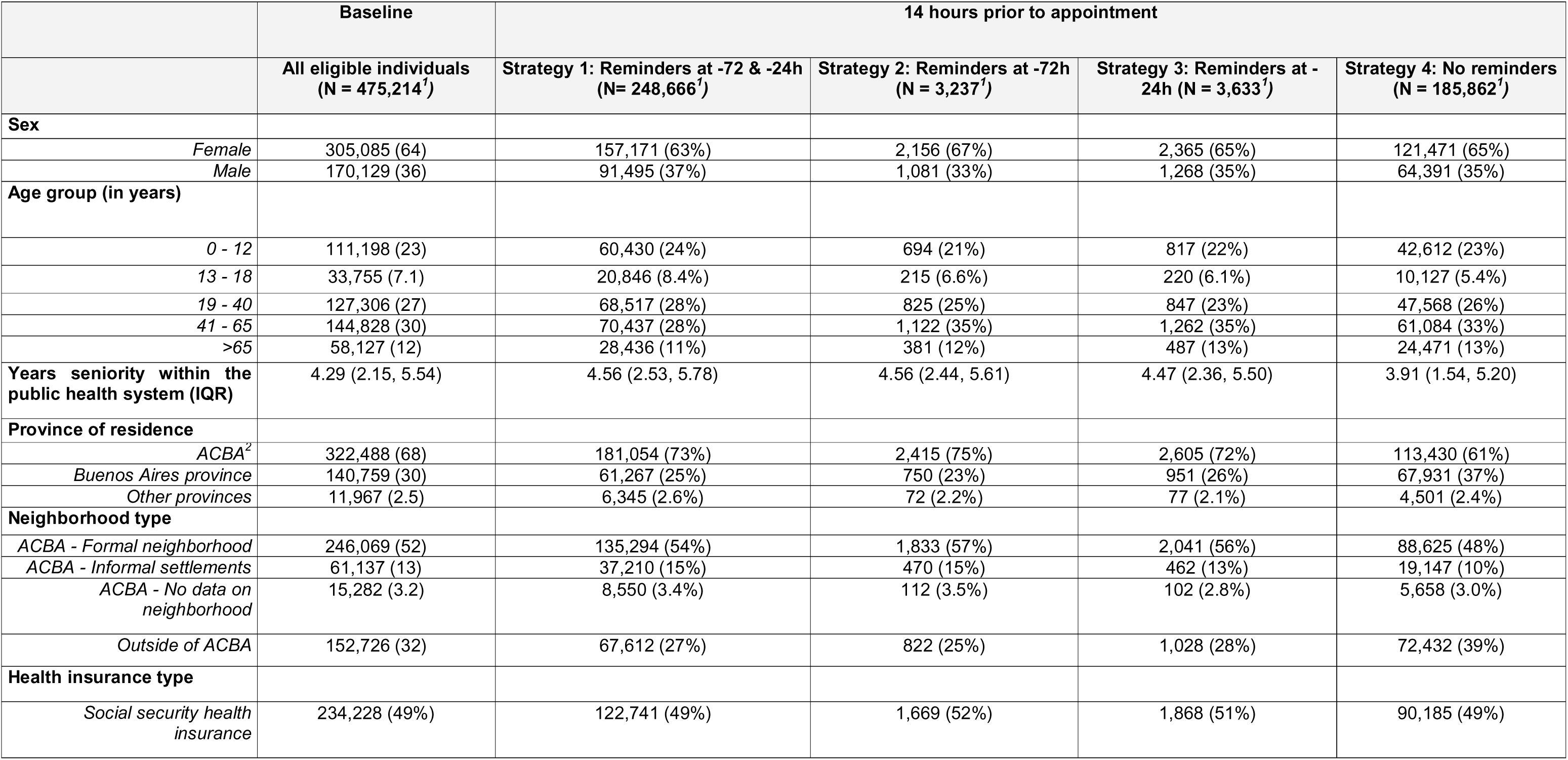
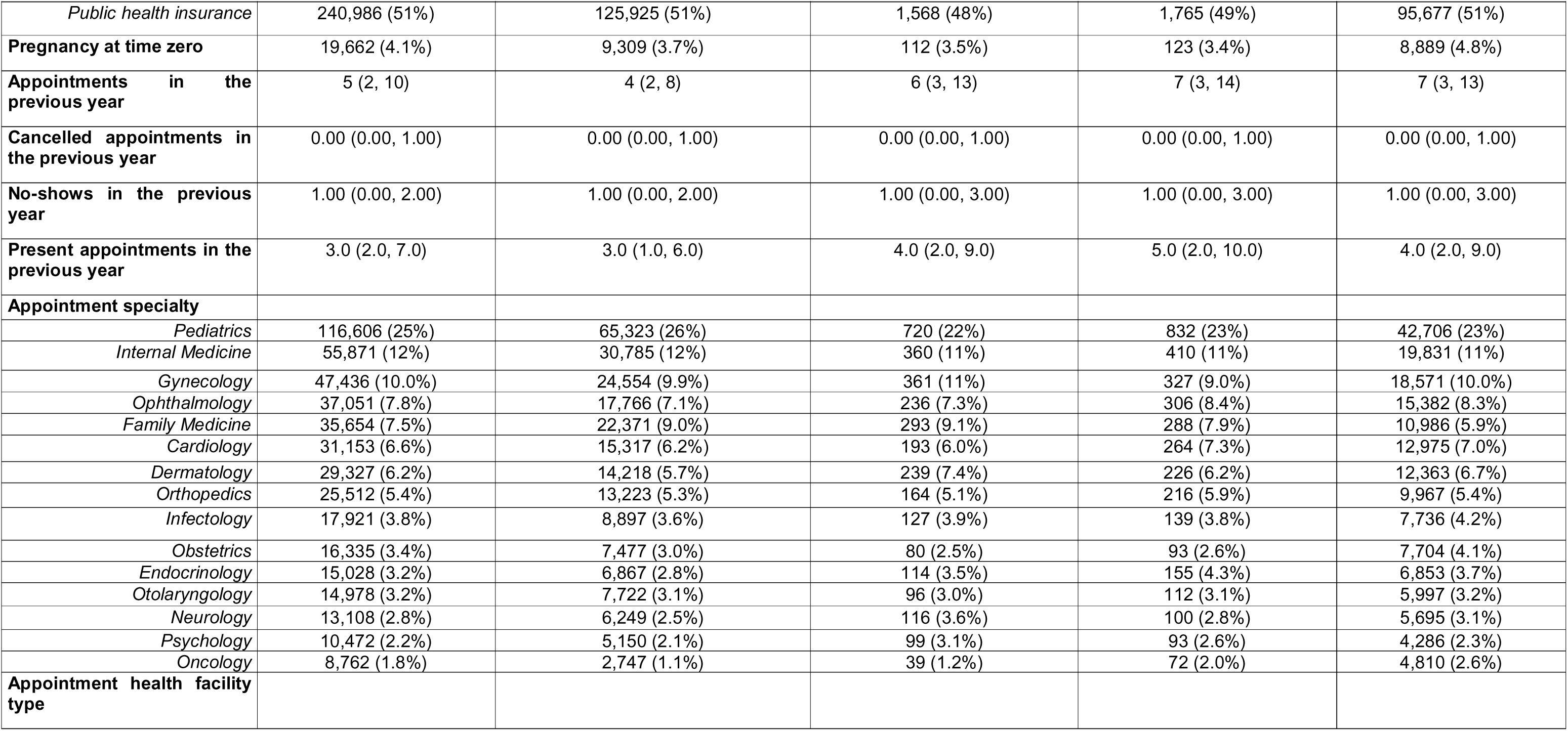
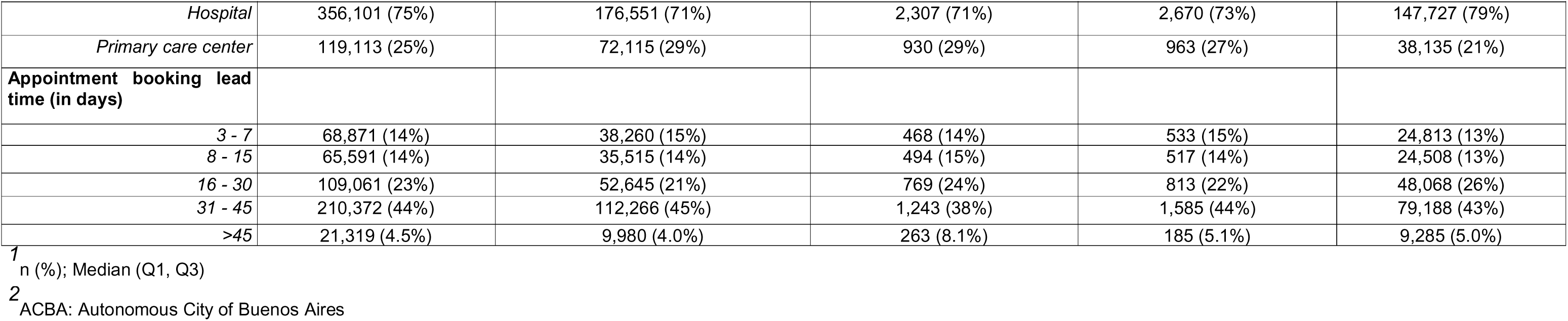
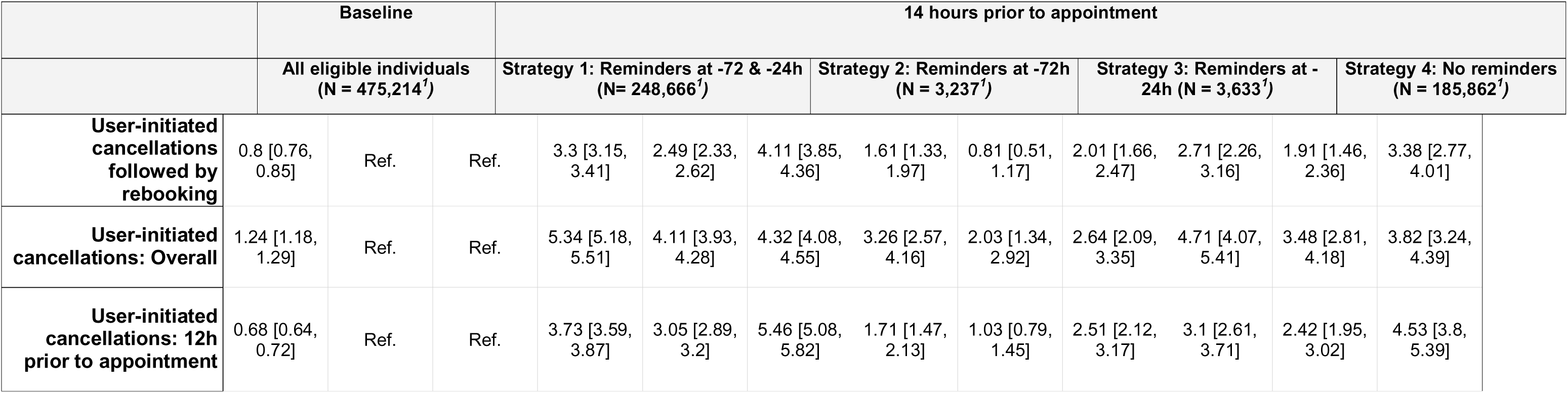
Selected baseline and 14 hours prior to appointment characteristics of eligible individuals for a target trial emulation of WhatsApp reminders and no-show cancellations, Buenos Aires Public Health System data (June 2023-May 2024). Numbers in parentheses are % unless otherwise specified.

The percentage of no-shows in each strategy was: 27.50% in the reminders 72 & 24h prior to appointment group (95% CI: 27.19, 27.81), 31.4% in the one reminder 72h prior to appointment group (95% CI: 28.87, 34.07), 26.34% in the one reminder 24h prior to appointment group (95% CI: 24.9, 27.93) and 34.55% in the no message group (95% CI: 34.33, 34.77). This corresponds to a risk difference of −7.05 percentage points (pp) [−7.41, −6.71] comparing the no message to the 72 & 24h reminders, −3.16pp [−5.69, −0.49] comparing the no message to the 72h reminder, and −8.21pp [−9.68, −6.54] comparing the no message to the 24h reminder (Table 3).

**Table 3.**
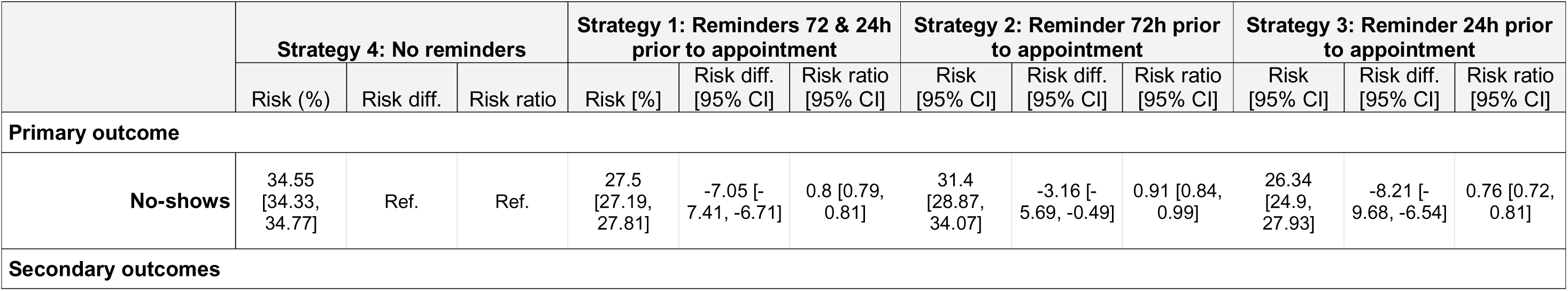
Estimated 75-hour risks of no shows (primary analysis), and user-initiated cancellations (secondary analysis) by treatment group, Buenos Aires Public Health System data (June 2023-May 2024).

Regarding secondary outcomes, the median follow-up period was 75 hours (interquartile range [IQR], 75 to 75). All reminder strategies led to an increase in user-initiated cancellations compared to the control group, including overall cancellations, late cancellations (within 12 hours), and cancellations followed by rebooking (Table 3, Figures 2 & 3). Overall user-initiated cancellations increased most notably with the two-reminder (RR: 4.32; RD: 4.11 pp) and 24-hour reminder (RR: 3.82; RD: 3.48 pp) strategies.

**Figure 2.**
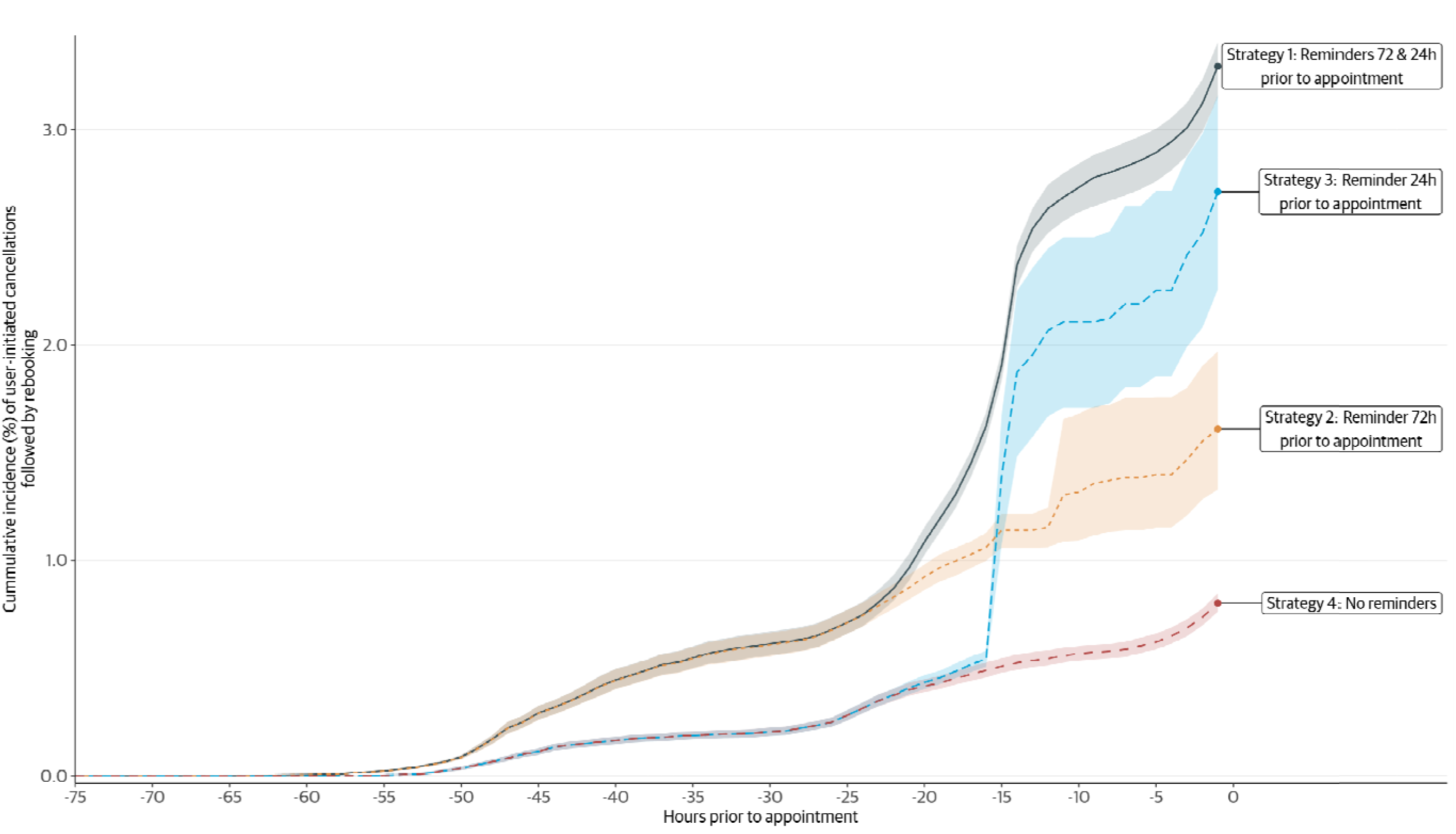
Estimated 75-hour risks of user-initiated cancellations followed by rebooking by treatment group, Buenos Aires Public Health System data (June 2023-May 2024)

**Figure 3.**
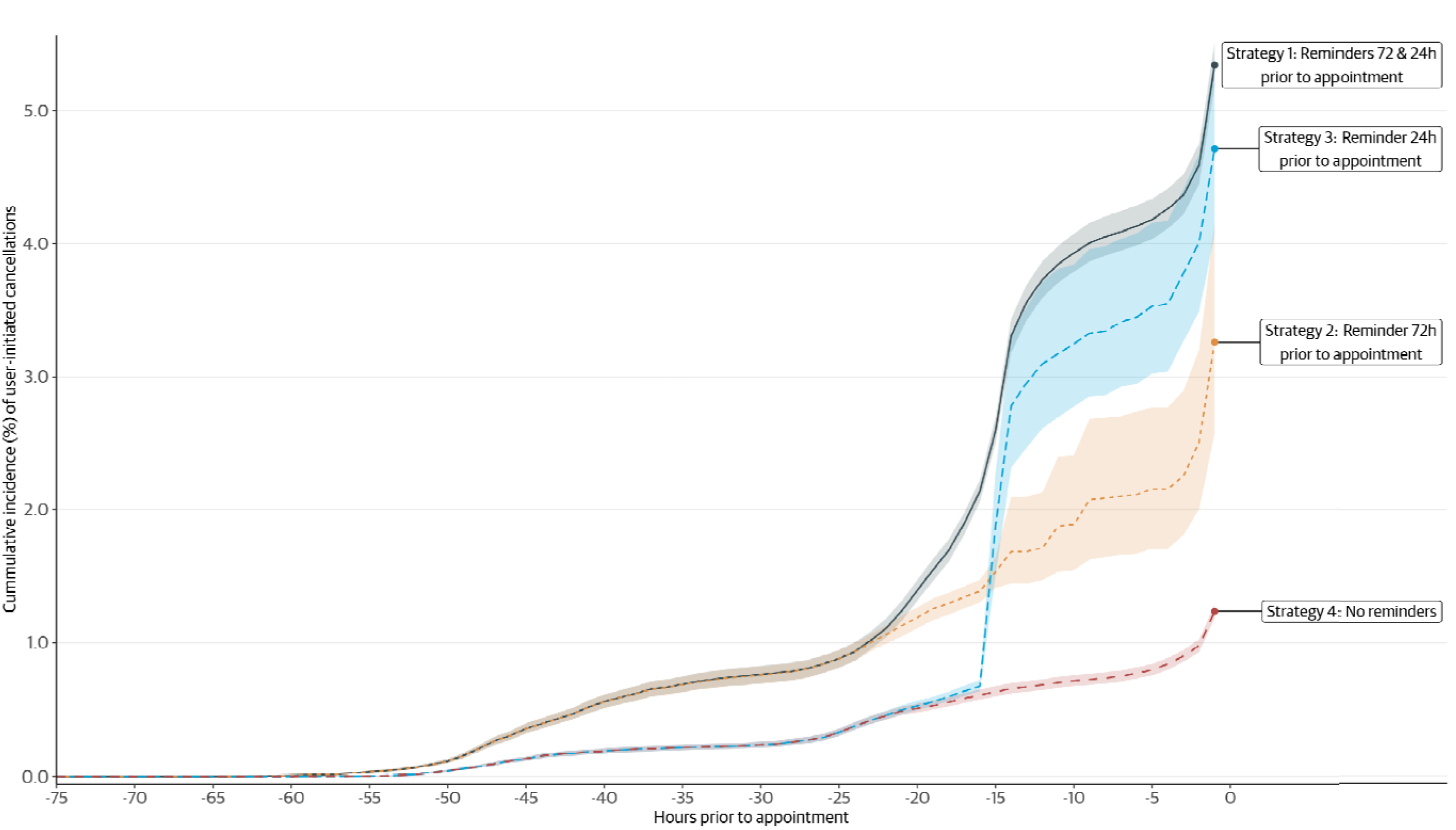
Estimated 75-hour risks of user-initiated cancellations treatment group, Buenos Aires Public Health System data (June 2023-May 2024)

Estimates were similar to those from the primary analysis within subgroups defined by health facility types, medical specialties, booking lead times, and age groups (Figure 4, Supplementary Material table 13). Notably, risk differences (were most pronounced in subgroups presenting with higher baseline no-show percentage.

**Figure 4.**
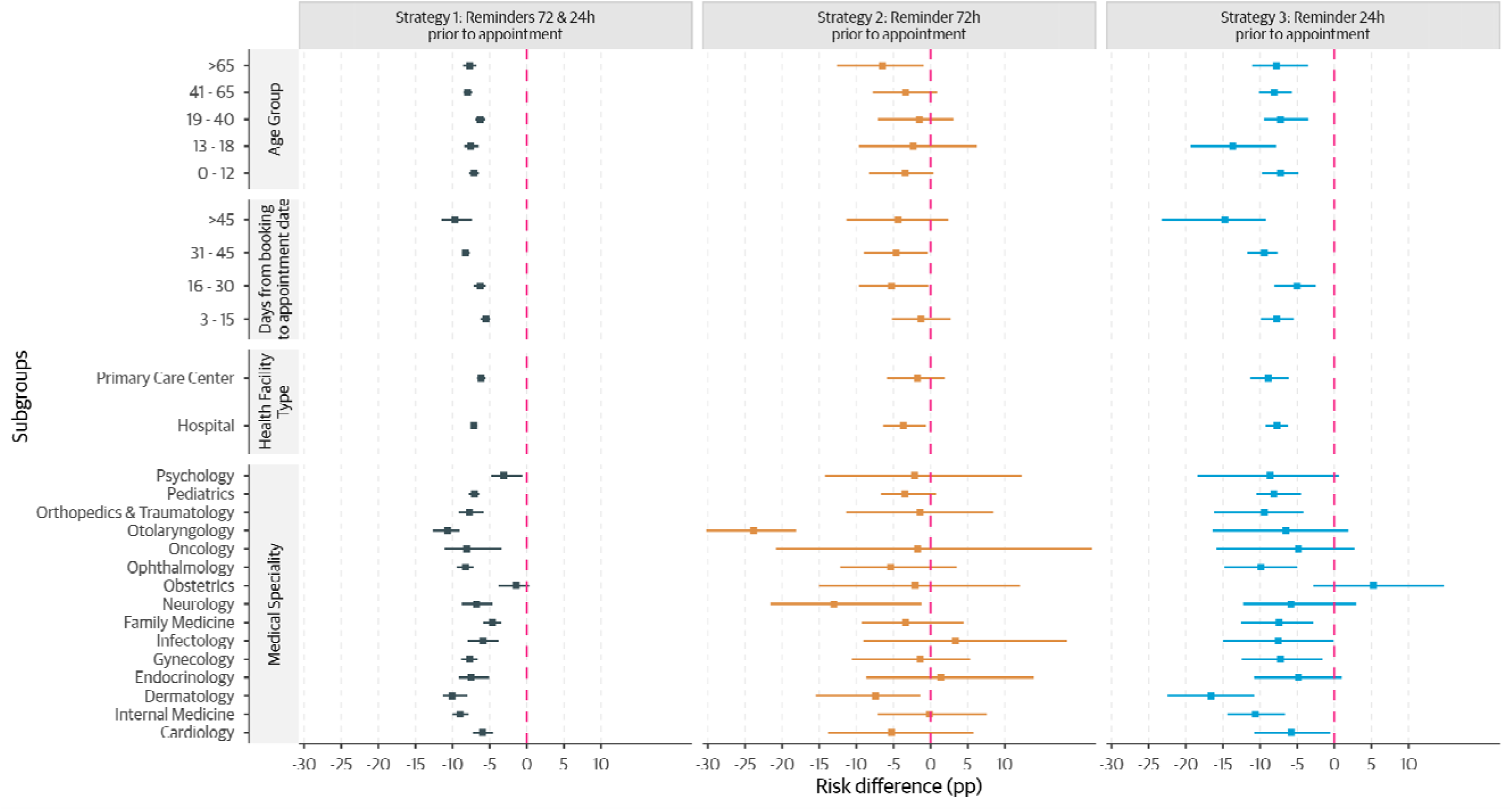
Estimated 75-hour risks of no-shows by treatment group, stratified by selected demographic and clinical characteristics, Buenos Aires Public Health System data (June 2023-May 2024)

Estimates were also consistent across sensitivity analyses (Supplementary Material Tables 14 & 15) and strategy effectiveness ranking was preserved, although the magnitude of some effect estimates was sensitive to assumptions regarding the timing of message delivery. Finally, the non-parametric estimates risks diverged from those obtained using the primary parametric pooled logistic regression model (Figure 4 of the Supplementary Material).

## Discussion

After emulating a target trial using a large public health system database from Buenos Aires, Argentina, we found that all three WhatsApp reminder strategies reduced the risk of no-shows compared to a strategy of no reminders. The single proximal (24-hour) reminder produced the largest absolute reduction (−8.21pp [−9.68, −6.54]; RR 0.76 [0.72, 0.81]), the dual-reminder strategy a comparable reduction (−7.05 pp [−7.41, −6.71]; RR 0.80 [0.79, 0.81]), and the single distal (72-hour) reminder a smaller, though still appreciable, reduction (−3.16 pp [−5.69, −0.49]; RR 0.91 [0.84, 0.99]). Taken together, these results indicate that timing, specifically the inclusion of a proximal cue, drives most of the effect, and that adding a second, more distal message yields no detectable additional benefit for attendance. Our results were similar after modifying several aspects of our study.

The relative-risk reductions observed for the two strategies that include a proximal cue (24% and 20% for the 24-hour-only and dual-reminder strategies, respectively) align with systematic reviews of SMS and other digital reminder interventions, which typically report reductions of approximately 25% in non-attendance[22,29]. The single 72-hour reminder produced a smaller reduction (9%) and falls below the range reported for proximal-reminder interventions in the existing literature. Our findings refine the previously suggested “dose-response” relationship between reminder frequency and attendance[30]: in our data, a single just-in-time nudge was at least as effective as a combined distal-plus-proximal schedule, while the distal-only schedule was clearly inferior.

The use of WhatsApp rather than a proprietary patient portal has practical advantages in this setting. WhatsApp is the dominant messaging platform in Latin America and is widely used in much of Africa and South and Southeast Asia, and embedding the intervention within an already-installed, familiar interface helps overcome the “app fatigue” associated with the proliferation of standalone mHealth applications[20,31]. The native bidirectional capabilities of the platform additionally allow patients to confirm or cancel within the same interface that delivered the reminder [32,33], which plausibly contributes to the observed increase in active cancellations. We caution, however, that this platform advantage is contingent on the underlying communication ecosystem: in settings where WhatsApp penetration is lower or where alternative messaging platforms dominate, the absolute benefit of a WhatsApp-based system may be smaller, and adaptation to the locally dominant channel would be required.

Absolute risk reductions tended to be largest in strata with the highest baseline no-show risk, including adolescents (13–18 years), appointments booked more than 45 days in advance, and certain specialties such as dermatology and internal medicine. These patterns suggest opportunities for targeting digital reminders toward populations and appointment types in which the baseline burden of absenteeism is greatest, and away from a uniform “one-size-fits-all” approach. However, the subgroup analyses were exploratory: 78 contrasts were estimated across 26 strata and three active strategies, without adjustment for multiple testing, and precision was limited in less prevalent strata and in the single 72-hour arm. Individual point estimates from these cells should be interpreted with caution and used to generate hypotheses for prospective evaluation rather than as definitive targets for differential intervention.

This study has limitations. First, eligibility for reminders relied on a proxy for mobile-phone ownership (number formatting), and we could not verify whether the message was read by the patient or by a proxy such as a relative; patients without registered mobile numbers, who are likely to be more socioeconomically vulnerable, are by design outside the population to which our estimates apply. Second, the cloning approach precludes an intention-to-treat analysis in the conventional sense, although in this context the per-protocol effect is arguably the more policy-relevant estimand. It isolates true efficacy from adherence failures[34,35], informing policymakers on whether it works as designed, not merely whether offering it shifts outcomes at the population level. If the PP effect substantially exceeds the ITT, adherence becomes a modifiable implementation target rather than evidence against the intervention itself.

Third, confounding is a key concern in any observational analysis because assignment of the reminder strategies was not randomized and thus the treatment groups may have different distributions of risk factors. For example, the no message group differed from other arms in geographic residence, facility type, and prior healthcare utilization. Beyond adjusting for all measured baseline covariates through inverse-probability-of-treatment weighting, we took several additional steps to address confounding: i) the study context itself provided partial protection against confounding by design: treatment assignment in the observed data was largely determined by the administrative availability and formatting accuracy of phone numbers in the Ministry of Health database, a factor that did not depend directly on patients’ clinical or behavioral characteristics, thereby approximating a natural experiment that limited systematic selection into reminder arms; ii) we assessed post-weighting balance empirically using standardized mean differences (SMDs) across all baseline covariates for each strategy compared to the control arm (Supplementary Material Figure 5), confirming that weighting achieved adequate covariate balance at the time of analysis.

Fourth, relatively few individuals remained protocol-adherent in the two single-reminder arms, only 3,237 in the 72-hour arm and 3,633 in the 24-hour arm, because the predominant real-world practice consisted of sending both reminders, leaving limited counterfactual contrast for the single-message strategies. As a result, estimates for these arms carry greater imprecision, particularly for the 72-hour strategy, whose confidence interval (RD −5.69 to −0.49 pp) is substantially wider than that of the dual-reminder strategy. We addressed this in two ways. First, we used non-parametric percentile bootstrap confidence intervals with 500 resamples, which accurately reflect the effective sample size without relying on asymptotic approximations that could underestimate uncertainty in small groups. Second, we pre-specified the sensitivity analysis varying the reminder-window cut-offs (Supplementary Material Table 14) to evaluate whether minor timing shifts changed the conclusions for these arms; the qualitative ranking of strategies was preserved across all specifications, although the 72-hour estimate showed greater sensitivity to window definition than the other strategies. These results should therefore be interpreted as providing directional evidence that a single distal reminder is less effective than a proximal or dual-reminder strategy, rather than as a precise point estimate of the magnitude of that difference.

Our results are important from a policy standpoint and may support the deployment of a single proximal WhatsApp reminder as the default strategy in resource-constrained outpatient systems: it was estimated to be at least as effective as a dual-reminder schedule for preventing no-shows, while potentially imposing a lower messaging cost on the system and a lower notification burden on patients. Where the operational goal extends beyond attendance to dynamic slot reallocation through active cancellations, a dual-reminder schedule may still be preferable, as it was the strongest driver of user-initiated cancellations in our data. Prospective studies designed to test targeted, adaptive reminder schedules in high-baseline-risk subgroups, and to evaluate the effect of these strategies in settings with different digital-communication ecosystems, would substantially strengthen the evidence base.

## Conclusion

In this large target trial emulation within the public health system of Buenos Aires, Argentina, automated WhatsApp reminders proved an effective intervention for reducing outpatient no-shows. Across all three active strategies, reminder delivery was associated with meaningful absolute reductions in absenteeism against a baseline no-show rate of 34.6%. Our findings refine the operational dose-response of digital reminders in two important ways. First, timing matters more than frequency: a single proximal reminder sent ∼24 hours before the appointment was clearly superior to a single distal reminder sent at 72 hours and was as effective as a dual-reminder schedule in preventing no-shows. Second, the dual-reminder strategy, while providing no additional benefit for attendance, was the strongest driver of user-initiated cancellations and may therefore retain operational value in settings where active slot reallocation is a priority. The consistency of results across subgroups and sensitivity analyses supports the robustness of these conclusions, notwithstanding the greater imprecision of the single-reminder arm estimates due to limited per-protocol adherence in the observed data. Taken together, these results support the deployment of a single just-in-time WhatsApp reminder as the default strategy in resource-constrained outpatient systems, and motivate prospective evaluation of targeted, adaptive reminder schedules in the high-baseline-risk subgroups identified here.

## Data Availability

The full, individual-level datasets are owned by Buenos Aires City Ministry of Health and are not publicly available due to data protection regulations. Interested researchers may request access to these data directly from Buenos Aires City Ministry of Health, subject to institutional and ethical approvals.

## Declarations

## Ethics approval and consent to participate

This study was approved by the Institutional Review Board at Hospital P. de Elizalde, under approval no. 15,103. Our research involved secondary analysis of identifiable data, accessed with appropriate institutional authorization and in compliance with national data protection laws. Strict measures were taken to ensure data confidentiality and secure storage. All necessary ethical safeguards were implemented, and the use of identifiable information was justified and approved by the ethics committee.

## Competing interests

The authors have declared that no competing interests exist

## Funding

This study was in part funded by a grant from the Inter-American Development Bank.

## Authors’ contributions

Protocol development: SE, GQ, AS; Conceptualization: SE, GQ, MS, AR, AS; Data curation: SE, GQ; Formal Analysis: SE, GQ; Investigation: SE, GQ; Methodology: SE, GQ, AS; Visualization: SE; Writing – original draft: SE; Writing, review & editing: SE, GQ, MS, AR, AS.

## Acknowledgements

Not applicable

## Notes

### Competing Interest Statement

The authors have declared no competing interest.

### Author Declarations

Institutional Review Board at Hospital P. de Elizalde gave ethical approval for this work

### Summary of Updates

We adjusted the estimated effect in the abstract; Added ORCID to one of the co-authors

