## Supplemental material for "Comparative Effectiveness of Single vs. Dual WhatsApp Reminders on No-shows: A Target Trial Emulation within the Public Health System of Buenos Aires, Argentina"

**Supplementary material**

**Implementation of a Whatsapp based chatbot for health appointment booking**

Since 2021, the Ministry of Health of the Autonomous City of Buenos Aires, Argentina, has undertaken a significant restructuring of the outpatient appointment scheduling process across its network of public hospitals and primary care centers^[1]^. This modernization required the development of new software and the management of major cultural and organizational shifts. A critical advancement occurred in early 2023 with the integration of "BOTI," the city's official WhatsApp chatbot^[2]^, into the scheduling system. This integration automated reminder delivery and facilitated appointment cancellations and rescheduling. The standard strategy involved sending two WhatsApp messages to each patient, approximately 72 and 24 hours prior to their appointment. These messages provided appointment details and offered a simple interface to cancel or reschedule at any time.

To receive WhatsApp reminders, patients were required to have a mobile phone number registered in the public healthcare database formatted compatible with the WhatsApp Application Programming Interface (API) used for message dispatch. However, during the initial rollout of the reminder strategy, significant variability in stored phone number formats was identified, which prevented the successful delivery of numerous messages. To address this limitation, a continuous data quality improvement initiative targeting phone numbers was implemented throughout 2023, gradually increasing the successful delivery rate over time.

Because the correct formatting of a mobile phone number in the Ministry of Health database was an administrative factor that did not depend directly on the patients’ Covariates, this context inadvertently created a natural experiment. Consequently, patients were heterogeneously exposed to one of the four intervention arms based solely on the availability and formatting accuracy of their contact data in the system, minimizing confounding bias.

In the scheduling system, if an appointment slot was booked, subsequently canceled by the patient, and then reassigned to a different individual, these events were recorded as two distinct booking instances. Furthermore, patients frequently scheduled more than one appointment during the study period (median 2 [IQR 1-3]). To accurately estimate the effect of initial exposure to the intervention, the primary analysis was restricted to a single appointment per patient, specifically, their first scheduled appointment within the study timeframe.

**Table 1 Suppl Material. Censoring criteria for protocol deviations in each arm and adherence probability for IP weights:**

- **Strategy 1: Two reminders, ~72 (75-64h) & ~24h (27-15h) prior to the appointment**

Individuals assigned to this strategy can be censored due to protocol deviation if:

1. Do not receive a reminder 64 hours prior to the appointment in the absence of a prior reminder between 75-65 hours
2. Do not receive a reminder 15 hours prior to the appointment in the absence of a prior reminder between 27 and 16 hours
3. User-initiated or healthcare system-initiated cancellation

| **Hours prior to appointment** | **Reminder sent: -75, -65h** | **Reminder sent: -64h** | **Reminder sent: -75, -64** | **Reminder sent: -63, -28h** | **Reminder sent: -27, -16h** | **Reminder sent: -15h** | **Reminder sent: -27, -15h** | **Adherence** | **Adherence probability** | **Method for adherence probability estimation** |
| --- | --- | --- | --- | --- | --- | --- | --- | --- | --- | --- |
| Between 65 and 75 | Any | - | - | - | - | - | - | Yes | 1 | Known by design |
| 64 | Yes | Any | Yes | - | - | - | - | Yes | 1 | Known by design |
|  | No | Yes | Yes | - | - | - | - | Yes | Prob. of receiving a reminder | **Model 1:** Prob. of receiving a reminder at -64h, when no reminder was received between -75 and -65h, conditional on baseline covariates |
|  |  | No | No | - | - | - | - | No | 0 | Censored observations have a weight of 0 |
| -63, -28h | Any | Any | Yes | Any | - | - | - | Yes | 1 | Known by design |
| -27, -16h | Any | Any | Yes | Any | Any | - | - | Yes | 1 | Known by design |
| -15h | Any | Any | Yes | Any | Yes | Any | Yes | Yes | 1 | Known by design |
|  |  |  |  |  | No | Yes | Yes | Yes | Prob. of receiving a reminder | **Model 2:** Prob. of receiving a reminder at -15h, when a reminder was received between -75 and -65h and no reminder was received between -27 and -16h, conditional on baseline covariates |
|  |  |  |  |  |  | No | No | No | 0 | Censored observations have a weight of 0 |
| -14, 0h | Any | Any | Yes | Any | Any | Any | Yes | Yes | 1 | Known by design |

**Table 2 Suppl Material. Models to estimate adherence probabilities in Strategy 1:**

| **Model #** | **Model for the…** | **Type of model** | **Baseline covariates** | **Person-hours** | **Number of person-hours (no. events)** |
| --- | --- | --- | --- | --- | --- |
| 1 | Probability of receiving a reminder | Logistic | - Sex (Female, Male)  - Days prior to appointment  - Total number of appointments in the previous year  - Total number of no-shows in the previous year  - Total number of present appointments in the previous year  - Health insurance (Yes, No)  - Type of neighborhood (ACBA formal, ACBA informal, Outside of ACBA, no data on neighborhood)  - Pregnancy (Yes, No)  - Type of health facility (primary care center, specialized ambulatory center, hospital)  - Medical specialty  - Week of time (cubic spline, 4 degrees of freedom)  - Age in years  - Total number of visits  -Seniority within the health system | 64 hours prior to appointment when no reminder was received between -75 and -65h | 245,697 (46,148) |
| 2 | Probability of receiving a reminder | Logistic | Same as above | 15 hours prior to appointment when a reminder was received between -75 and -65h and no reminder was received between -27 and -16h | 64,358 (48,393) |
| 3 | System initiated cancellation | Logistic | - Hour (cubic spline, 6 degrees of freedom)  - Sex (Female, Male)  - Days prior to appointment  - Total number of appointments in the previous year  - Total number of no-shows in the previous year  - Total number of present appointments in the previous year  - Health insurance (Yes, No)  - Type of neighborhood (ACBA formal, ACBA informal, Outside of ACBA, no data on neighborhood)  - Pregnancy (Yes, No)  - Type of health facility (primary care center, specialized ambulatory center, hospital)  - Medical specialty  - Week of time (cubic spline, 4 degrees of freedom)  - Age in years  - Total number of visits  -Seniority within the health system | All hours | 35,206,873 (9,130) |

| **Model 1: Model to estimate the probability of receiving a reminder 64 hours prior to the appointment** | | | |  | **Model 2: Model to estimate the probability of receiving a reminder 15 hours prior to the appointment** | | | |
| --- | --- | --- | --- | --- | --- | --- | --- | --- |
| **Covariate** | **Beta** | **SE** | **p-value** |  | **Covariate** | **Beta** | **SE** | **p-value** |
| **Sex** |  |  |  |  | **Sex** |  |  |  |
| *Female* | — | — |  |  | *Female* | — | — |  |
| *Male* | 0.009 | 0.03 | 0.770 |  | *Male* | 0.073 | 0.05 | 0.111 |
| **Age** | 0.002 | 0.00 | 0.044 |  | **Age** | -0.002 | 0.00 | 0.228 |
| **Days prior to appointment** | 0.002 | 0.00 | 0.033 |  | **Days prior to appointment** | -0.005 | 0.00 | <0.001 |
| **# appointments last year** | 0.027 | 0.01 | 0.044 |  | **# appointments last year** | -0.024 | 0.02 | 0.210 |
| **# no-shows last year** | -0.059 | 0.02 | <0.001 |  | **# no-shows last year** | 0.014 | 0.02 | 0.536 |
| **# present appointments last year** | -0.041 | 0.01 | 0.006 |  | **# present appointments last year** | 0.027 | 0.02 | 0.182 |
| **Health insurance type** |  |  |  |  | **Health insurance type** |  |  |  |
| *Public health insurance* | — | — |  |  | *Public health insurance* | — | — |  |
| *Social security* | 0.093 | 0.03 | 0.002 |  | *Social security* | 0.030 | 0.04 | 0.476 |
| **Neighborhood type** |  |  |  |  | **Neighborhood type** |  |  |  |
| *ACBA formal* | — | — |  |  | *ACBA formal* | — | — |  |
| *ACBA informal* | 0.072 | 0.03 | 0.037 |  | *ACBA informal* | 0.097 | 0.05 | 0.049 |
| *No data on neighborhood* | -0.243 | 0.06 | <0.001 |  | *No data on neighborhood* | 0.013 | 0.09 | 0.888 |
| *Outside of ACBA* | -0.464 | 0.05 | <0.001 |  | *Outside of ACBA* | 0.014 | 0.07 | 0.850 |
| **Pregnancy** |  |  |  |  | **Pregnancy** |  |  |  |
| *No* | — | — |  |  | *No* | — | — |  |
| *Yes* | 0.217 | 0.09 | 0.015 |  | *Yes* | -0.353 | 0.12 | 0.005 |
| **Appointment health facility type** |  |  |  |  | **Appointment health facility type** |  |  |  |
| *Primary care center* | — | — |  |  | *Primary care center* | — | — |  |
| *Specialized ambulatory center* | 0.401 | 0.14 | 0.004 |  | *Specialized ambulatory center* | -0.117 | 0.19 | 0.534 |
| **Appointment health specialty** |  |  |  |  | **Appointment health specialty** |  |  |  |
| *Cardiology* | — | — |  |  | *Cardiology* | — | — |  |
| *Dermatology* | -0.002 | 0.24 | 0.994 |  | *Dermatology* | 0.376 | 0.34 | 0.273 |
| *Endocrinology* | -0.439 | 0.28 | 0.117 |  | *Endocrinology* | 0.385 | 0.40 | 0.340 |
| *Family Medicine* | -0.066 | 0.18 | 0.710 |  | *Family Medicine* | 0.316 | 0.23 | 0.172 |
| *Gynecology* | 0.514 | 0.18 | 0.004 |  | *Gynecology* | 0.224 | 0.24 | 0.344 |
| *Infectology* | -12.27 | 101 | 0.904 |  | *Infectology* | 10.88 | 97 | 0.910 |
| *Internal Medicine* | 0.604 | 0.18 | <0.001 |  | *Internal Medicine* | 0.233 | 0.24 | 0.322 |
| *Neurology* | 0.224 | 0.31 | 0.468 |  | *Neurology* | 0.469 | 0.45 | 0.296 |
| *Obstetrics* | 0.448 | 0.21 | 0.035 |  | *Obstetrics* | 1.025 | 0.29 | <0.001 |
| *Ophthalmology* | -0.779 | 0.32 | 0.014 |  | *Ophthalmology* | -0.026 | 0.43 | 0.951 |
| *Orthopedics* | 0.716 | 0.35 | 0.044 |  | *Orthopedics* | -0.298 | 0.47 | 0.530 |
| *Otolaryngology* | 0.117 | 0.43 | 0.785 |  | *Otolaryngology* | -0.256 | 0.60 | 0.672 |
| *Pediatrics* | 0.468 | 0.18 | 0.010 |  | *Pediatrics* | 0.217 | 0.24 | 0.367 |
| *Psychology* | 0.689 | 0.19 | <0.001 |  | *Psychology* | 0.110 | 0.25 | 0.659 |
| **splines::ns(`Week of time zero`, df = 4)** |  |  |  |  | **splines::ns(`Week of time zero`, df = 4)** |  |  |  |
| *splines::ns(`Week of time zero`, df = 4)1* | 1.369 | 0.08 | <0.001 |  | *splines::ns(`Week of time zero`, df = 4)1* | 1.257 | 0.10 | <0.001 |
| *splines::ns(`Week of time zero`, df = 4)2* | 2.942 | 0.07 | <0.001 |  | *splines::ns(`Week of time zero`, df = 4)2* | 0.428 | 0.11 | <0.001 |
| *splines::ns(`Week of time zero`, df = 4)3* | 0.873 | 0.17 | <0.001 |  | *splines::ns(`Week of time zero`, df = 4)3* | 0.878 | 0.18 | <0.001 |
| *splines::ns(`Week of time zero`, df = 4)4* | 2.757 | 0.07 | <0.001 |  | *splines::ns(`Week of time zero`, df = 4)4* | 1.186 | 0.11 | <0.001 |
| **# visits within the health system** | 0.003 | 0.00 | 0.040 |  | **# visits within the health system** | -0.002 | 0.00 | 0.393 |
| **Seniority within the health system** | 0.169 | 0.01 | <0.001 |  | **Seniority within the health system** | 0.017 | 0.01 | 0.103 |

Abbreviation: SE = Standard Error

| **Model 3: Model to estimate the probability of a system-initiated cancellation** | | | |
| --- | --- | --- | --- |
| **Covariate** | **Beta** | **SE** | **p-value** |
| **splines::ns(Hour, df = 6)** |  |  |  |
| *splines::ns(Hour, df = 6)1* | -54.39 | 20 | 0.008 |
| *splines::ns(Hour, df = 6)2* | -47.20 | 21 | 0.022 |
| *splines::ns(Hour, df = 6)3* | -53.72 | 20 | 0.009 |
| *splines::ns(Hour, df = 6)4* | -37.54 | 15 | 0.011 |
| *splines::ns(Hour, df = 6)5* | -90.27 | 38 | 0.018 |
| *splines::ns(Hour, df = 6)6* | -28.27 | 8.6 | <0.001 |
| **Sex** |  |  |  |
| *Female* | — | — |  |
| *Male* | 0.045 | 0.04 | 0.216 |
| **Age** | -0.001 | 0.00 | 0.589 |
| **Days prior to appointment** | 0.010 | 0.00 | <0.001 |
| **# appointments last year** | -0.017 | 0.01 | 0.260 |
| **# no-shows last year** | 0.031 | 0.02 | 0.090 |
| **# present appointments last year** | 0.017 | 0.02 | 0.293 |
| **Health insurance type** |  |  |  |
| *Public health insurance* | — | — |  |
| *Social security* | 0.029 | 0.03 | 0.400 |
| **Neighborhood type** |  |  |  |
| *ACBA formal* | — | — |  |
| *ACBA informal* | 0.038 | 0.04 | 0.337 |
| *No data on neighborhood* | -0.025 | 0.07 | 0.721 |
| *Outside of ACBA* | -0.019 | 0.06 | 0.737 |
| **Pregnancy** |  |  |  |
| *No* | — | — |  |
| *Yes* | 0.251 | 0.11 | 0.021 |
| **Appointment health facility type** |  |  |  |
| *Primary care center* | — | — |  |
| *Specialized ambulatory center* | -0.140 | 0.17 | 0.397 |
| **Appointment health specialty** |  |  |  |
| *Cardiology* | — | — |  |
| *Dermatology* | -0.764 | 0.25 | 0.002 |
| *Endocrinology* | 0.599 | 0.39 | 0.123 |
| *Family Medicine* | -0.238 | 0.22 | 0.274 |
| *Gynecology* | -0.548 | 0.22 | 0.013 |
| *Infectology* | 1.809 | 1.0 | 0.076 |
| *Internal Medicine* | -0.096 | 0.22 | 0.667 |
| *Neurology* | -0.009 | 0.41 | 0.983 |
| *Obstetrics* | 0.111 | 0.27 | 0.678 |
| *Ophthalmology* | 0.557 | 0.41 | 0.171 |
| *Orthopedics* | 1.654 | 1.0 | 0.105 |
| *Otolaryngology* | 1.582 | 1.0 | 0.121 |
| *Pediatrics* | -0.228 | 0.22 | 0.307 |
| *Psychology* | -0.094 | 0.24 | 0.690 |
| **splines::ns(`Week of time zero`, df = 4)** |  |  |  |
| *splines::ns(`Week of time zero`, df = 4)1* | -0.011 | 0.08 | 0.897 |
| *splines::ns(`Week of time zero`, df = 4)2* | 0.566 | 0.09 | <0.001 |
| *splines::ns(`Week of time zero`, df = 4)3* | -0.005 | 0.15 | 0.972 |
| *splines::ns(`Week of time zero`, df = 4)4* | -0.796 | 0.07 | <0.001 |
| **# visits within the health system** | -0.001 | 0.00 | 0.422 |
| **Seniority within the health system** | -0.001 | 0.01 | 0.885 |
| Abbreviations: SE = Standard Error | | |  |

**Table 3 Suppl Material. IP Weights distribution before 99th percentile truncation**

| **Reminder strategy** | **Mean** | **p1** | **p99** | **Minimum** | **Maximum** | **Range** |
| --- | --- | --- | --- | --- | --- | --- |
| Strategy 1 | 1.638098 | 1 | 18.20416 | 1 | 277.0891 | 276.0891 |

**Table 4 Suppl Material. Censoring criteria for protocol deviations in each arm and adherence probability for IP weights:**

**Strategy 2: One reminder, ~72 (75-64h) prior to the appointment**

**Individuals assigned to this strategy are censored if:**

- Do not receive a reminder 64 hours prior to the appointment in the absence of a prior reminder between 75-65 hours
- Receive a reminder at any time after that
- User-initiated or healthcare system-initiated cancellation

| **Hours prior to appointment time** | **Reminder sent: -75, -65h** | **Reminder sent: -64h** | **Reminder sent: -75, -64** | **Reminder sent: -63, -28h** | **Reminder sent: -27, -15h** | **Adherence** | **Adherence probability** | **Method for adherence probability estimation** |
| --- | --- | --- | --- | --- | --- | --- | --- | --- |
| -75, -65h | Any | - | - | - | - | Yes | 1 | Known by design |
| -64h | Yes | Any | Yes | - | - | Yes | 1 | Known by design |
|  | No | Yes | Yes | - | - | Yes | Prob. of receiving a reminder at -64h, when no reminder was received between -75 and -65h | **Model 1:** Prob. of receiving a reminder at -64h, when no reminder was received between -75 and -65h conditional on baseline covariates |
|  |  | No | No | - | - | No | 0 | Censored observations receive a weight of 0 |
| -63, -28h | Any | Any | Yes | Any | - | Yes | 1 | Known by design |
| -27, -15h | Any | Any | Yes | Any | No | Yes | 1 - (Prob. of receiving a reminder between -27 and -15h, when a reminder was received between -75 and -64h) | **Model 4**: Prob. of receiving a reminder between -27 and -15h, when a reminder was received between -75 and -64h conditional on baseline covariates |
| -14, 0h | Any | Any | Yes | Any | Yes | Yes | 1 | Known by design |

**Table 5 Suppl Material. Models to estimate adherence probabilities in Strategy 2:**

| **Model #** | **Model for the…** | **Type of model** | **Baseline covariates** | **Person-hours** | **Number of person-hours (no. events)** |
| --- | --- | --- | --- | --- | --- |
| 4 | Probability of receiving a reminder | Logistic | - Hour (cubic spline, 6 degrees of freedom)  - Sex (Female, Male)  - Days prior to appointment  - Total number of appointments in the previous year  - Total number of no-shows in the previous year  - Total number of present appointments in the previous year  - Health insurance (Yes, No)  - Type of neighborhood (ACBA formal, ACBA informal, Outside of ACBA, no data on neighborhood)  - Pregnancy (Yes, No)  - Type of health facility (primary care center, specialized ambulatory center, hospital)  - Medical specialty  - Week of time (cubic spline, 4 degrees of freedom)  - Age in years  - Total number of visits  -Seniority within the health system | Between -27 and -15h, when a reminder was received between -75 and -64h | 3,506,271 (253,632) |

**Health system-initiated cancellation model same as above.**

| **Model 4: Model to estimate the probability of receiving a reminder 27 -15 hours prior to the appointment** | | | |
| --- | --- | --- | --- |
| **Covariate** | **Beta** | **SE** | **p-value** |
| **splines::ns(Hour, df = 6)** |  |  |  |
| *splines::ns(Hour, df = 6)1* | 25.33 | 1.6 | <0.001 |
| *splines::ns(Hour, df = 6)2* | 28.04 | 1.6 | <0.001 |
| *splines::ns(Hour, df = 6)3* | 26.37 | 1.6 | <0.001 |
| *splines::ns(Hour, df = 6)4* | 20.68 | 1.1 | <0.001 |
| *splines::ns(Hour, df = 6)5* | 48.73 | 3.1 | <0.001 |
| *splines::ns(Hour, df = 6)6* | 13.27 | 0.66 | <0.001 |
| **Sex** |  |  |  |
| *Female* | — | — |  |
| *Male* | 0.000 | 0.01 | 0.994 |
| **Age** | 0.000 | 0.00 | 0.524 |
| **Days prior to appointment** | 0.000 | 0.00 | 0.068 |
| **# appointments last year** | 0.000 | 0.00 | 0.978 |
| **# no-shows last year** | 0.000 | 0.00 | 0.921 |
| **# present appointments last year** | 0.000 | 0.00 | 0.951 |
| **Health insurance type** |  |  |  |
| *Public health insurance* | — | — |  |
| *Social security* | 0.005 | 0.01 | 0.508 |
| **Neighborhood type** |  |  |  |
| *ACBA formal* | — | — |  |
| *ACBA informal* | 0.003 | 0.01 | 0.754 |
| *No data on neighborhood* | 0.003 | 0.02 | 0.879 |
| *Outside of ACBA* | -0.004 | 0.01 | 0.795 |
| **Pregnancy** |  |  |  |
| *No* | — | — |  |
| *Yes* | -0.027 | 0.03 | 0.283 |
| **Appointment health facility type** |  |  |  |
| *Primary care center* | — | — |  |
| *Specialized ambulatory center* | -0.026 | 0.05 | 0.592 |
| **Appointment health specialty** |  |  |  |
| *Cardiology* | — | — |  |
| *Dermatology* | 0.017 | 0.08 | 0.828 |
| *Endocrinology* | 0.033 | 0.09 | 0.709 |
| *Family Medicine* | 0.033 | 0.06 | 0.565 |
| *Gynecology* | 0.004 | 0.06 | 0.946 |
| *Infectology* | 0.089 | 0.12 | 0.445 |
| *Internal Medicine* | 0.005 | 0.06 | 0.937 |
| *Neurology* | 0.003 | 0.11 | 0.980 |
| *Obstetrics* | 0.030 | 0.07 | 0.652 |
| *Ophthalmology* | 0.043 | 0.09 | 0.616 |
| *Orthopedics* | -0.044 | 0.13 | 0.734 |
| *Otolaryngology* | 0.010 | 0.13 | 0.937 |
| *Pediatrics* | 0.001 | 0.06 | 0.985 |
| *Psychology* | -0.009 | 0.06 | 0.880 |
| **splines::ns(`Week of time zero`, df = 4)** |  |  |  |
| *splines::ns(`Week of time zero`, df = 4)1* | 0.073 | 0.02 | <0.001 |
| *splines::ns(`Week of time zero`, df = 4)2* | 0.036 | 0.02 | 0.093 |
| *splines::ns(`Week of time zero`, df = 4)3* | 0.078 | 0.04 | 0.048 |
| *splines::ns(`Week of time zero`, df = 4)4* | 0.056 | 0.02 | 0.005 |
| **# visits within the health system** | 0.000 | 0.00 | 0.691 |
| **Seniority within the health system** | -0.001 | 0.00 | 0.738 |
| Abbreviation: OR = Odds Ratio | | |  |

**Table 6 Suppl Material. IP Weights distribution before 99th percentile truncation**

| **Reminder strategy** | **Mean** | **p1** | **p99** | **Minimum** | **Maximum** | **Range** |
| --- | --- | --- | --- | --- | --- | --- |
| Strategy 2 | 2.225778 | 1 | 25.5788 | 1 | 366.6732 | 365.6732 |

**Table 7 Suppl Material. Censoring criteria for protocol deviations in each arm and adherence probability for IP weights:**

**Strategy 3: One reminder, ~24h (27-15h) prior to the appointment**

**Individuals assigned to this strategy are censored if:**

- Receive a reminder in the first window: between 75-64 hours prior to the appointment
- Do not receive a reminder 15 hours prior to the appointment in the absence of a prior reminder between 27 and 16 hours
- User-initiated or healthcare system-initiated cancellation

| **Hours prior to appointment time** | **Reminder sent: -75, -64** | **Reminder sent: -63, -28h** | **Reminder sent: -27, -16h** | **Reminder sent: -15h** | **Reminder sent: -27, -15h** | **Adherence** | **Adherence probability** | **Method for adherence probability estimation** |
| --- | --- | --- | --- | --- | --- | --- | --- | --- |
| -75, -64h | No | - | - | - | - | Yes | 1 - (Prob. of receiving a reminder between -75 and -64h) | **Model 5:** Prob. of receiving a reminder between -75 and -64h, conditional on baseline covariates |
|  | Yes | - | - | - | - | No | 0 | Censored observations receive a weight of 0 |
| -63, -28h | No | Any | - | - | - | Yes | 1 | Known by design |
| -27, -16h | No | Any | Any | - | - | Yes | 1 | Known by design |
| -15h | No | Any | Yes | Any | Yes | Yes | 1 | Known by design |
|  |  |  | No | Yes | Yes | Yes | Prob. of receiving a reminder at -15h, when no reminder was received in the past | **Model 6:** Prob. of receiving a reminder at -15h, when no reminder was received in the past, conditional on baseline covariates |
|  |  |  |  | No | No | No | 0 | Known by design |
| -14, 0h | Yes | Any | Any | Any | Yes | Yes | 1 | Known by design |

**Models to estimate adherence probabilities in Strategy 3:**

| **Model #** | **Model for the** | **Type of model** | **Baseline covariates** | **Person-hours** | **Number of person-hours (no. events)** |
| --- | --- | --- | --- | --- | --- |
| 5 | Probability of receiving a reminder | Logistic | Same as above | Between -75 and -64h | 3,307,979 (275,665) |
| 6 | Probability of receiving a reminder | Logistic | Same as above | Hour 15 when no prior reminder was received | 187,038 (833) |

**Health system-initiated cancellation model same as above.**

| **Model 5: Model to estimate the probability of receiving a reminder 75-64 hours prior to the appointment** | | | |
| --- | --- | --- | --- |
| **Covariate** | **Beta** | **SE** | **p-value** |
| **splines::ns(Hour, df = 6)** |  |  |  |
| *splines::ns(Hour, df = 6)1* | 5.720 | 0.15 | <0.001 |
| *splines::ns(Hour, df = 6)2* | 7.149 | 0.16 | <0.001 |
| *splines::ns(Hour, df = 6)3* | 5.584 | 0.15 | <0.001 |
| *splines::ns(Hour, df = 6)4* | 6.058 | 0.10 | <0.001 |
| *splines::ns(Hour, df = 6)5* | 10.58 | 0.31 | <0.001 |
| *splines::ns(Hour, df = 6)6* | 3.893 | 0.06 | <0.001 |
| **Sex** |  |  |  |
| *Female* | — | — |  |
| *Male* | 0.000 | 0.01 | >0.999 |
| **Age** | 0.000 | 0.00 | >0.999 |
| **Days prior to appointment** | 0.000 | 0.00 | >0.999 |
| **# appointments last year** | 0.000 | 0.00 | >0.999 |
| **# no-shows last year** | 0.000 | 0.00 | >0.999 |
| **# present appointments last year** | 0.000 | 0.00 | >0.999 |
| **Health insurance type** |  |  |  |
| *Public health insurance* | — | — |  |
| *Social security* | 0.000 | 0.01 | >0.999 |
| **Neighborhood type** |  |  |  |
| *ACBA formal* | — | — |  |
| *ACBA informal* | 0.000 | 0.01 | >0.999 |
| *No data on neighborhood* | 0.000 | 0.02 | >0.999 |
| *Outside of ACBA* | 0.000 | 0.01 | >0.999 |
| **Pregnancy** |  |  |  |
| *No* | — | — |  |
| *Yes* | 0.000 | 0.02 | >0.999 |
| **Appointment health facility type** |  |  |  |
| *Primary care center* | — | — |  |
| *Specialized ambulatory center* | 0.000 | 0.05 | >0.999 |
| **Appointment health specialty** |  |  |  |
| *Cardiology* | — | — |  |
| *Dermatology* | 0.000 | 0.08 | >0.999 |
| *Endocrinology* | 0.000 | 0.08 | >0.999 |
| *Family Medicine* | 0.000 | 0.06 | >0.999 |
| *Gynecology* | 0.000 | 0.06 | >0.999 |
| *Infectology* | 0.000 | 0.11 | >0.999 |
| *Internal Medicine* | 0.000 | 0.06 | >0.999 |
| *Neurology* | 0.000 | 0.10 | >0.999 |
| *Obstetrics* | 0.000 | 0.06 | >0.999 |
| *Ophthalmology* | 0.000 | 0.08 | >0.999 |
| *Orthopedics* | 0.000 | 0.12 | >0.999 |
| *Otolaryngology* | 0.000 | 0.12 | >0.999 |
| *Pediatrics* | 0.000 | 0.06 | >0.999 |
| *Psychology* | 0.000 | 0.06 | >0.999 |
| **splines::ns(`Week of time zero`, df = 4)** |  |  |  |
| *splines::ns(`Week of time zero`, df = 4)1* | 0.000 | 0.02 | >0.999 |
| *splines::ns(`Week of time zero`, df = 4)2* | 0.000 | 0.02 | >0.999 |
| *splines::ns(`Week of time zero`, df = 4)3* | 0.000 | 0.04 | >0.999 |
| *splines::ns(`Week of time zero`, df = 4)4* | 0.000 | 0.02 | >0.999 |
| **# visits within the health system** | 0.000 | 0.00 | >0.999 |
| **Seniority within the health system** | 0.000 | 0.00 | >0.999 |

| **Model 6: Model to estimate the probability of receiving a reminder 15 hours prior to the appointment** | | | |
| --- | --- | --- | --- |
| **Covariate** | **Beta** | **SE** | **p-value** |
| **Sex** |  |  |  |
| *Female* | — | — |  |
| *Male* | 0.287 | 0.17 | 0.094 |
| **Age** | 0.000 | 0.01 | 0.981 |
| **Days prior to appointment** | 0.000 | 0.00 | 0.920 |
| **# appointments last year** | 0.019 | 0.06 | 0.743 |
| **# no-shows last year** | -0.017 | 0.07 | 0.798 |
| **# present appointments last year** | -0.029 | 0.06 | 0.640 |
| **Health insurance type** |  |  |  |
| *Public health insurance* | — | — |  |
| *Social security* | 0.073 | 0.16 | 0.658 |
| **Neighborhood type** |  |  |  |
| *ACBA formal* | — | — |  |
| *ACBA informal* | -0.214 | 0.19 | 0.272 |
| *No data on neighborhood* | -0.885 | 0.46 | 0.056 |
| *Outside of ACBA* | -0.450 | 0.30 | 0.132 |
| **Pregnancy** |  |  |  |
| *No* | — | — |  |
| *Yes* | -0.205 | 0.56 | 0.713 |
| **Appointment health facility type** |  |  |  |
| *Primary care center* | — | — |  |
| *Specialized ambulatory center* | -1.082 | 0.90 | 0.231 |
| **Appointment health specialty** |  |  |  |
| *Cardiology* | — | — |  |
| *Dermatology* | 1.121 | 1.2 | 0.366 |
| *Endocrinology* | -11.90 | 348 | 0.973 |
| *Family Medicine* | -0.292 | 1.1 | 0.786 |
| *Gynecology* | 0.137 | 1.1 | 0.900 |
| *Infectology* | -12.06 | 761 | 0.987 |
| *Internal Medicine* | 0.133 | 1.1 | 0.902 |
| *Neurology* | 2.714 | 1.3 | 0.035 |
| *Obstetrics* | 0.831 | 1.2 | 0.498 |
| *Ophthalmology* | -12.32 | 470 | 0.979 |
| *Orthopedics* | 2.625 | 1.5 | 0.086 |
| *Otolaryngology* | -11.92 | 914 | 0.990 |
| *Pediatrics* | 0.138 | 1.1 | 0.900 |
| *Psychology* | 1.265 | 1.1 | 0.252 |
| **splines::ns(`Week of time zero`, df = 4)** |  |  |  |
| *splines::ns(`Week of time zero`, df = 4)1* | 1.049 | 0.34 | 0.002 |
| *splines::ns(`Week of time zero`, df = 4)2* | -0.127 | 0.53 | 0.810 |
| *splines::ns(`Week of time zero`, df = 4)3* | -1.577 | 0.82 | 0.055 |
| *splines::ns(`Week of time zero`, df = 4)4* | -0.299 | 0.64 | 0.640 |
| **# visits within the health system** | 0.003 | 0.01 | 0.560 |
| **Seniority within the health system** | 0.059 | 0.04 | 0.143 |

**Table 8 Suppl Material. IP Weights distribution before 99th percentile truncation**

| **Reminder strategy** | **Mean** | **p1** | **p99** | **Minimum** | **Maximum** | **Range** |
| --- | --- | --- | --- | --- | --- | --- |
| Strategy 3 | 1.757379 | 1.002427 | 2.801911 | 1.000405 | 1970.114 | 1969.113 |

**Table 9 Suppl Material. Censoring criteria for protocol deviations in each arm and adherence probability for IP weights:**

**Strategy 4: No reminders prior to the appointment**

| **Hours prior to appointment time** | **Reminder sent: -75, 0h** | **Adherence** | **Adherence probability** | **Method for adherence probability estimation** |
| --- | --- | --- | --- | --- |
| -75, -0h | No | Yes | 1 - (Prob. of receiving a reminder between -75 and 0h) | Model 7: Prob. of receiving a reminder between -75 and 0h |
|  | Yes | No | 0 | Censored observations receive a weight of 0 |

| **Model #** | **Model for the** | **Type of model** | **Baseline covariates** | **Person-hours** | **Number of person-hours (no. events)** |
| --- | --- | --- | --- | --- | --- |
| 7 | Probability of receiving a reminder | Logistic | Same as above | Between -75 and 0 | 35,655,306 (286,387) |

| **Model 7: Model to estimate the probability of receiving a reminder prior to the appointment** | | | |
| --- | --- | --- | --- |
| **Covariate** | **Beta** | **SE** | **p-value** |
| **splines::ns(Hour, df = 6)** |  |  |  |
| *splines::ns(Hour, df = 6)1* | -24.13 | 0.14 | <0.001 |
| *splines::ns(Hour, df = 6)2* | -16.55 | 2.6 | <0.001 |
| *splines::ns(Hour, df = 6)3* | -3.392 | 0.60 | <0.001 |
| *splines::ns(Hour, df = 6)4* | 0.693 | 0.57 | 0.221 |
| *splines::ns(Hour, df = 6)5* | -10.59 | 1.3 | <0.001 |
| *splines::ns(Hour, df = 6)6* | -33.18 | 2.3 | <0.001 |
| **Sex** |  |  |  |
| *Female* | — | — |  |
| *Male* | -0.002 | 0.01 | 0.788 |
| **Age** | 0.000 | 0.00 | 0.175 |
| **Days prior to appointment** | 0.000 | 0.00 | 0.143 |
| **# appointments last year** | 0.000 | 0.00 | 0.991 |
| **# no-shows last year** | -0.007 | 0.00 | 0.138 |
| **# present appointments last year** | -0.006 | 0.00 | 0.112 |
| **Health insurance type** |  |  |  |
| *Public health insurance* | — | — |  |
| *Social security* | 0.039 | 0.01 | <0.001 |
| **Neighborhood type** |  |  |  |
| *ACBA formal* | — | — |  |
| *ACBA informal* | 0.010 | 0.01 | 0.266 |
| *No data on neighborhood* | -0.046 | 0.02 | 0.005 |
| *Outside of ACBA* | -0.123 | 0.01 | <0.001 |
| **Pregnancy** |  |  |  |
| *No* | — | — |  |
| *Yes* | -0.006 | 0.02 | 0.808 |
| **Appointment health facility type** |  |  |  |
| *Primary care center* | — | — |  |
| *Specialized ambulatory center* | -0.063 | 0.04 | 0.158 |
| **Appointment health specialty** |  |  |  |
| *Cardiology* | — | — |  |
| *Dermatology* | 0.039 | 0.07 | 0.584 |
| *Endocrinology* | -0.026 | 0.08 | 0.746 |
| *Family Medicine* | 0.092 | 0.05 | 0.084 |
| *Gynecology* | 0.064 | 0.05 | 0.239 |
| *Infectology* | 0.162 | 0.11 | 0.140 |
| *Internal Medicine* | 0.095 | 0.05 | 0.080 |
| *Neurology* | 0.147 | 0.10 | 0.138 |
| *Obstetrics* | 0.133 | 0.06 | 0.030 |
| *Ophthalmology* | 0.093 | 0.08 | 0.241 |
| *Orthopedics* | 0.137 | 0.12 | 0.247 |
| *Otolaryngology* | 0.095 | 0.12 | 0.423 |
| *Pediatrics* | 0.055 | 0.05 | 0.316 |
| *Psychology* | 0.315 | 0.06 | <0.001 |
| **splines::ns(`Week of time zero`, df = 4)** |  |  |  |
| *splines::ns(`Week of time zero`, df = 4)1* | 1.387 | 0.02 | <0.001 |
| *splines::ns(`Week of time zero`, df = 4)2* | 0.823 | 0.02 | <0.001 |
| *splines::ns(`Week of time zero`, df = 4)3* | 1.184 | 0.04 | <0.001 |
| *splines::ns(`Week of time zero`, df = 4)4* | 1.026 | 0.02 | <0.001 |
| **# visits within the health system** | 0.001 | 0.00 | 0.016 |
| **Seniority within the health system** | 0.036 | 0.00 | <0.001 |

**Table 10 Suppl Material. IP Weights distribution before 99th percentile truncation**

| **Reminder strategy** | **Mean** | **p1** | **p99** | **Minimum** | **Maximum** | **Range** |
| --- | --- | --- | --- | --- | --- | --- |
| Strategy 4 | 1.455775 | 1.000921 | 2.767986 | 1.000137 | 4.61943 | 3.619293 |

**Table 11 Suppl Material. Follow-up, cancellations, adherence and no-shows per intervention strategy.**

| **Intervention strategy** | **Follow-up (hours) [Median (IQR)]** | **Healthcare system-initiated cancellations (censored)**  **n (%)** | **Non-adherent (censored)**  **n (%)** | **User-initiated cancellations**  **n (%)** | **Attended the appointment**  **n (%)** | **No-shows**  **n (%)** |
| --- | --- | --- | --- | --- | --- | --- |
| **Strategy 1 (Reminders ~72 & ~24h prior to appointment)** | 75 (11-75) | 5,472 (1.15) | 215,492 (45.35) | 14,200 (2.99) | 240,050 (50.51) | 72,038 (15.16) |
| **Strategy 2 (Reminder ~72 prior to appointment)** | 54 (11-58) | 2,670 (0.56) | 464,022 (97.64) | 5,394 (1.14) | 3,128 (0.66) | 989 (0.21) |
| **Strategy 3 (Reminder ~24h prior to appointment)** | 11 (8-60) | 1,801 (0.38) | 468,431 (98.57) | 1,492 (0.31) | 3,490 (0.73) | 957 (0.20) |
| **Strategy 4 (No reminders)** | 11 (8-75) | 3,442 (0.72) | 286,308 (60.25) | 2,392 (0.50)) | 183,072 (38.52) | 63,423 (13.35) |

**Table 12. Contrast between active strategies for no-shows**

|  | **Outcome** | **Risk diff. (pp)[95% CI]** | **Risk ratio [95% CI]** |
| --- | --- | --- | --- |
| Strategy 3: Reminder 24h prior to appointment  vs. Strategy 1: Reminders 72 & 24h prior to appointment | No-shows | -1.16 [-2.7, 0.44] | 0.96 [0.9, 1.02] |
| Strategy 3: Reminder 24h prior to appointment  vs. Strategy 2: Reminder 72h prior to appointment | No-shows | -5.06 [-7.83, -2.14] | 0.84 [0.77, 0.93] |
| Strategy 1: Reminders 72 & 24h prior to appointment  vs. Strategy 2: Reminder 72h prior to appointment | No-shows | -3.9 [-6.6, -1.32] | 0.88 [0.81, 0.95] |

| **Table 13 Suppl Material. Subgroup analysis for no-shows.** | | | | | | | | | | | | |
| --- | --- | --- | --- | --- | --- | --- | --- | --- | --- | --- | --- | --- |
|  | **Strategy 4: No reminders** | | | **Strategy 1: Reminders 72 & 24h prior to appointment** | | | **Strategy 2: Reminder 72h prior to appointment** | | | **Strategy 3: Reminder 24h prior to appointment** | | |
|  | **CI (%)** | **Risk diff. (pp)** | **Risk ratio** | **CI (%)** | **Risk diff. (pp)** | **Risk ratio** | **CI (%)** | **Risk diff. (pp)** | **Risk ratio** | **CI (%)** | **Risk diff. (pp)** | **Risk ratio** |
| **Health Facility Type** | | | | | | | | | | | | |
| **Hospital** | 35.36 [35.07, 35.59] | Ref. | Ref. | 28.21 [27.84, 28.57] | -7.15 [-7.5, -6.7] | 0.8 [0.79, 0.81] | 31.69 [28.91, 34.66] | -3.67 [-6.4, -0.67] | 0.9 [0.82, 0.98] | 27.63 [26.08, 29.06] | -7.74 [-9.27, -6.27] | 0.78 [0.74, 0.82] |
| **Primary care center** | 31.54 [31.13, 31.95] | Ref. | Ref. | 25.35 [24.97, 25.89] | -6.19 [-6.7, -5.53] | 0.8 [0.79, 0.82] | 29.79 [25.18, 33.44] | -1.76 [-5.86, 1.92] | 0.94 [0.81, 1.06] | 22.64 [20.22, 25.45] | -8.9 [-11.33, -6.13] | 0.72 [0.64, 0.81] |
| **Appointment specialty** | | | | | | | | | | | | |
| **Cardiology** | 28.82 [28.14, 29.52] | Ref. | Ref. | 22.87 [21.38, 24.17] | -5.95 [-7.27, -4.52] | 0.79 [0.75, 0.84] | 23.57 [15.18, 34.19] | -5.25 [-13.82, 5.78] | 0.82 [0.52, 1.2] | 23 [17.65, 28.13] | -5.83 [-10.8, -0.53] | 0.8 [0.62, 0.98] |
| **Internal Medicine** | 40.77 [40.01, 41.57] | Ref. | Ref. | 31.81 [31.03, 32.58] | -8.96 [-10.02, -7.83] | 0.78 [0.76, 0.81] | 40.58 [33.28, 48.34] | -0.18 [-7.17, 7.58] | 1 [0.82, 1.19] | 30.12 [26.15, 34.33] | -10.65 [-14.36, -6.65] | 0.74 [0.65, 0.84] |
| **Dermatology** | 43.37 [42.32, 44.18] | Ref. | Ref. | 33.3 [32.23, 34.75] | -10.07 [-11.29, -8.02] | 0.77 [0.74, 0.81] | 35.97 [28.05, 42.23] | -7.4 [-15.45, -1.33] | 0.83 [0.64, 0.97] | 26.77 [20.72, 32.48] | -16.6 [-22.46, -10.79] | 0.62 [0.48, 0.75] |
| **Endocrinology** | 24.79 [23.69, 25.66] | Ref. | Ref. | 17.29 [16.05, 18.77] | -7.51 [-9.17, -5.08] | 0.7 [0.64, 0.79] | 26.18 [16.11, 38.83] | 1.39 [-8.69, 13.9] | 1.06 [0.65, 1.56] | 19.95 [13.81, 25.47] | -4.85 [-10.82, 0.99] | 0.8 [0.56, 1.04] |
| **Gynecology** | 36.84 [36.24, 37.45] | Ref. | Ref. | 29.13 [28.09, 30.1] | -7.71 [-8.82, -6.62] | 0.79 [0.76, 0.82] | 35.43 [26.69, 42.21] | -1.41 [-10.63, 5.38] | 0.96 [0.72, 1.15] | 29.59 [24.58, 35.38] | -7.25 [-12.46, -1.61] | 0.8 [0.66, 0.96] |
| **Infectology** | 37.09 [36.15, 38.34] | Ref. | Ref. | 31.2 [29.51, 33.18] | -5.89 [-7.97, -3.8] | 0.84 [0.79, 0.9] | 40.39 [28.18, 55.26] | 3.3 [-9.01, 18.35] | 1.09 [0.76, 1.5] | 29.54 [22.46, 36.99] | -7.55 [-14.99, -0.08] | 0.8 [0.6, 1] |
| **Family Medicine** | 33.18 [32.24, 34.11] | Ref. | Ref. | 28.56 [27.95, 29.17] | -4.62 [-5.89, -3.42] | 0.86 [0.83, 0.89] | 29.79 [24.17, 37.3] | -3.39 [-9.27, 4.47] | 0.9 [0.72, 1.13] | 25.73 [20.87, 30.57] | -7.45 [-12.57, -2.83] | 0.78 [0.63, 0.91] |
| **Neurology** | 31.46 [30.32, 32.63] | Ref. | Ref. | 24.68 [22.99, 26.36] | -6.78 [-8.77, -4.62] | 0.78 [0.73, 0.85] | 18.47 [9.98, 29.74] | -12.99 [-21.56, -1.19] | 0.59 [0.32, 0.96] | 25.61 [19.2, 34.28] | -5.85 [-12.26, 2.95] | 0.81 [0.61, 1.09] |
| **Obstetrics** | 29.9 [28.94, 30.74] | Ref. | Ref. | 28.44 [26.27, 29.82] | -1.46 [-3.8, 0.34] | 0.95 [0.87, 1.01] | 27.8 [14.98, 41.66] | -2.1 [-15.03, 12.06] | 0.93 [0.5, 1.41] | 35.17 [26.99, 44.12] | 5.27 [-2.82, 14.68] | 1.18 [0.91, 1.5] |
| **Ophthalmology** | 31.88 [31.29, 32.64] | Ref. | Ref. | 23.64 [22.61, 24.61] | -8.24 [-9.42, -7.17] | 0.74 [0.71, 0.77] | 26.51 [19.78, 35.79] | -5.37 [-12.16, 3.52] | 0.83 [0.62, 1.11] | 21.98 [17.14, 26.97] | -9.9 [-14.78, -4.99] | 0.69 [0.54, 0.84] |
| **Oncology** | 26.57 [25.48, 27.67] | Ref. | Ref. | 18.46 [15.54, 22.58] | -8.1 [-11.07, -3.4] | 0.7 [0.58, 0.87] | 24.83 [5.69, 48.1] | -1.73 [-20.84, 21.74] | 0.93 [0.22, 1.84] | 21.72 [11.31, 29.39] | -4.85 [-15.9, 2.72] | 0.82 [0.42, 1.1] |
| **Otolaryngology** | 40.51 [39.36, 42.01] | Ref. | Ref. | 29.85 [28.49, 31.46] | -10.66 [-12.68, -9.05] | 0.74 [0.7, 0.77] | 16.68 [10.3, 21.83] | -23.84 [-30.19, -18.08] | 0.41 [0.26, 0.54] | 33.98 [24.46, 42.95] | -6.53 [-16.39, 1.91] | 0.84 [0.6, 1.05] |
| **Orthopedics & Traumatology** | 35.74 [34.83, 36.57] | Ref. | Ref. | 27.99 [26.96, 29.4] | -7.74 [-9.18, -5.82] | 0.78 [0.75, 0.83] | 34.32 [25.01, 44.41] | -1.42 [-11.32, 8.45] | 0.96 [0.69, 1.24] | 26.3 [18.73, 31.34] | -9.43 [-16.2, -4.17] | 0.74 [0.54, 0.88] |
| **Pediatrics** | 32.8 [32.3, 33.27] | Ref. | Ref. | 25.75 [25.18, 26.19] | -7.05 [-7.82, -6.38] | 0.79 [0.76, 0.8] | 29.3 [26.06, 33.3] | -3.49 [-6.67, 0.74] | 0.89 [0.8, 1.02] | 24.66 [22.29, 28.26] | -8.13 [-10.47, -4.49] | 0.75 [0.68, 0.86] |
| **Psychology** | 38.86 [37.59, 40.41] | Ref. | Ref. | 35.76 [33.78, 37.89] | -3.11 [-4.8, -0.6] | 0.92 [0.88, 0.98] | 36.7 [25.32, 50.37] | -2.17 [-14.21, 12.27] | 0.94 [0.64, 1.32] | 30.21 [20.73, 39.54] | -8.65 [-18.44, 0.64] | 0.78 [0.53, 1.02] |
| **Appointment booking lead time (in days)** | | | | | | | | | | | | |
| **3 - 15** | 29.87 [29.59, 30.34] | Ref. | Ref. | 24.37 [23.89, 24.89] | -5.5 [-6.25, -4.94] | 0.82 [0.79, 0.83] | 28.54 [24.83, 32.91] | -1.32 [-5.2, 2.65] | 0.96 [0.83, 1.09] | 22.08 [20, 24.37] | -7.79 [-9.89, -5.49] | 0.74 [0.67, 0.82] |
| **16 - 30** | 33.52 [33.13, 33.93] | Ref. | Ref. | 27.25 [26.46, 27.96] | -6.27 [-7.16, -5.51] | 0.81 [0.79, 0.83] | 28.25 [23.95, 33.5] | -5.26 [-9.68, -0.29] | 0.84 [0.71, 0.99] | 28.48 [25.48, 31.14] | -5.04 [-8.07, -2.51] | 0.85 [0.76, 0.93] |
| **31 - 45** | 37.08 [36.74, 37.34] | Ref. | Ref. | 28.82 [28.43, 29.26] | -8.26 [-8.71, -7.68] | 0.78 [0.77, 0.79] | 32.42 [28.18, 36.58] | -4.66 [-8.95, -0.37] | 0.87 [0.76, 0.99] | 27.64 [25.56, 29.54] | -9.44 [-11.73, -7.64] | 0.75 [0.69, 0.79] |
| **>45** | 43.9 [42.84, 44.85] | Ref. | Ref. | 34.22 [32.61, 35.95] | -9.69 [-11.5, -7.39] | 0.78 [0.74, 0.83] | 39.51 [32.45, 46.38] | -4.39 [-11.29, 2.41] | 0.9 [0.74, 1.05] | 29.17 [20.79, 34.42] | -14.73 [-23.23, -9.23] | 0.66 [0.47, 0.79] |
| **Age Group (in years)** | | | | | | | | | | | | |
| **0 - 12** | 32.94 [32.46, 33.33] | Ref. | Ref. | 25.8 [25.23, 26.36] | -7.13 [-7.76, -6.45] | 0.78 [0.77, 0.8] | 29.48 [24.64, 33.03] | -3.45 [-8.3, 0.36] | 0.9 [0.75, 1.01] | 25.71 [22.97, 28.11] | -7.22 [-9.76, -4.84] | 0.78 [0.7, 0.85] |
| **13 - 18** | 35.39 [34.49, 36.13] | Ref. | Ref. | 27.83 [27.08, 28.54] | -7.56 [-8.41, -6.48] | 0.79 [0.77, 0.81] | 33.04 [25.34, 41.34] | -2.35 [-9.69, 6.23] | 0.93 [0.73, 1.18] | 21.72 [16.02, 27.22] | -13.67 [-19.35, -7.85] | 0.61 [0.45, 0.77] |
| **19 - 40** | 40.92 [40.47, 41.34] | Ref. | Ref. | 34.65 [34.05, 35.27] | -6.28 [-6.92, -5.59] | 0.85 [0.83, 0.86] | 39.41 [33.83, 44.13] | -1.51 [-7.12, 3.1] | 0.96 [0.83, 1.08] | 33.68 [31.45, 37.37] | -7.24 [-9.46, -3.52] | 0.82 [0.77, 0.91] |
| **41 - 65** | 33.16 [32.83, 33.48] | Ref. | Ref. | 25.19 [24.7, 25.78] | -7.97 [-8.5, -7.35] | 0.76 [0.74, 0.78] | 29.78 [25.29, 33.88] | -3.39 [-7.79, 0.89] | 0.9 [0.76, 1.03] | 25.06 [22.95, 27.24] | -8.1 [-10.15, -5.71] | 0.76 [0.69, 0.83] |
| **>65** | 27.89 [27.29, 28.46] | Ref. | Ref. | 20.18 [19.42, 21.15] | -7.71 [-8.6, -6.77] | 0.72 [0.7, 0.76] | 21.41 [15.1, 26.96] | -6.48 [-12.59, -0.92] | 0.77 [0.55, 0.97] | 20.08 [16.82, 24.11] | -7.81 [-11.03, -3.54] | 0.72 [0.6, 0.87] |

| **Table 14 Suppl Material. Sensitivity Analysis:** Cut-off point used to assign each participant to a reminder strategy (+- 2h for each reminder window), primary outcome. | | | | | | | | | | | | |
| --- | --- | --- | --- | --- | --- | --- | --- | --- | --- | --- | --- | --- |
|  | **Strategy 4: No reminders** | | | **Strategy 1: Reminders 72 & 24h prior to appointment** | | | **Strategy 2: Reminder 72h prior to appointment** | | | **Strategy 3: Reminder 24h prior to appointment** | | |
| **Reminder window  cut-off strategy** | **CI (%)** | **Risk diff. (pp)** | **Risk ratio** | **CI (%)** | **Risk diff. (pp)** | **Risk ratio** | **CI (%)** | **Risk diff. (pp)** | **Risk ratio** | **CI (%)** | **Risk diff. (pp)** | **Risk ratio** |
| Primary analysis  (75-64h, 27-15h) | 34.55 [34.33, 34.77] | Ref. | Ref. | 27.5 [27.19, 27.81] | -7.05 [-7.41, -6.71] | 0.8 [0.79, 0.81] | 31.4 [28.87, 34.07] | -3.16 [-5.69, -0.49] | 0.91 [0.84, 0.99] | 26.34 [24.9, 27.93] | -8.21 [-9.68, -6.54] | 0.76 [0.72, 0.81] |
| Narrower 1st & 2nd  (75-66h, 27-17h) | 34.55 | Ref. | Ref. | 27.72 | -6.84 | 0.8 | 28.46* | -6.09* | 0.82* | 26.31 | -8.24 | 0.76 |
| Narrower 1st  (75-66h, 27-15h) | 34.55 | Ref. | Ref. | 27.8 | -6.75 | 0.8 | 28.46* | -6.09* | 0.82* | 26.34 | -8.21 | 0.76 |
| Narrower 2nd  (75-64h, 27-17h) | 34.55 | Ref. | Ref. | 28.59* | -5.97* | 0.83* | 31.4 | -3.16 | 0.91 | 26.31 | -8.24 | 0.76 |
| Wider 1st & 2nd  (75-62h, 27-13h) | 34.55 | Ref. | Ref. | 28.35* | -6.2* | 0.82* | 30.57 | -3.99 | 0.88 | 27 | -7.55 | 0.78 |
| Wider 1st, narrower 2nd  (75-62h, 27-17h) | 34.55 | Ref. | Ref. | 28.54* | -6.01* | 0.83* | 30.57 | -3.99 | 0.88 | 26.31 | -8.24 | 0.76 |
| Wider 1st  (75-62h, 27-15h) | 34.55 | Ref. | Ref. | 28.31* | -6.24* | 0.82* | 30.57 | -3.99 | 0.88 | 26.34 | -8.21 | 0.76 |
| Wider 2nd  (75-64h, 27-13h) | 34.55 | Ref. | Ref. | 27.56 | -6.99 | 0.8 | 31.4 | -3.16 | 0.91 | 27 | -7.55 | 0.78 |
| * Value is outside of the 95% CI | | | | | | | | | | | | |

| **Table 15 Suppl Material. Sensitivity Analysis:** Modelling strategies including interactions. | | | | | | | | | | | | |
| --- | --- | --- | --- | --- | --- | --- | --- | --- | --- | --- | --- | --- |
|  | **Strategy 4: No reminders** | | | **Strategy 1: Reminders 72 & 24h prior to appointment** | | | **Strategy 2: Reminder 72h prior to appointment** | | | **Strategy 3: Reminder 24h prior to appointment** | | |
| **Modelling strategy** | **CI (%)** | **Risk diff. (pp)** | **Risk ratio** | **CI (%)** | **Risk diff. (pp)** | **Risk ratio** | **CI (%)** | **Risk diff. (pp)** | **Risk ratio** | **CI (%)** | **Risk diff. (pp)** | **Risk ratio** |
| Primary analysis | 34.55 [34.33, 34.77] | Ref. | Ref. | 27.5 [27.19, 27.81] | -7.05 [-7.41, -6.71] | 0.8 [0.79, 0.81] | 31.4 [28.87, 34.07] | -3.16 [-5.69, -0.49] | 0.91 [0.84, 0.99] | 26.34 [24.9, 27.93] | -8.21 [-9.68, -6.54] | 0.76 [0.72, 0.81] |
| Primary analysis + interactions:  gender, age and medical specialty | 34.56 | Ref. | Ref. | 27.51 | -7.06 | 0.8 | 31.4 | -3.16 | 0.91 | 26.34 | -8.22 | 0.76 |
| Primary analysis + interactions:  gender and age | 34.56 | Ref. | Ref. | 27.51 | -7.06 | 0.8 | 31.4 | -3.16 | 0.91 | 26.34 | -8.22 | 0.76 |
| Primary analysis + interactions:  gender and medical specialty | 34.56 | Ref. | Ref. | 27.51 | -7.06 | 0.8 | 31.4 | -3.16 | 0.91 | 26.34 | -8.22 | 0.76 |
| Primary analysis + interactions:  age and medical specialty | 34.56 | Ref. | Ref. | 27.51 | -7.06 | 0.8 | 31.4 | -3.16 | 0.91 | 26.34 | -8.22 | 0.76 |
| * Value is outside of the 95% CI | | | | | | | | | | | | |

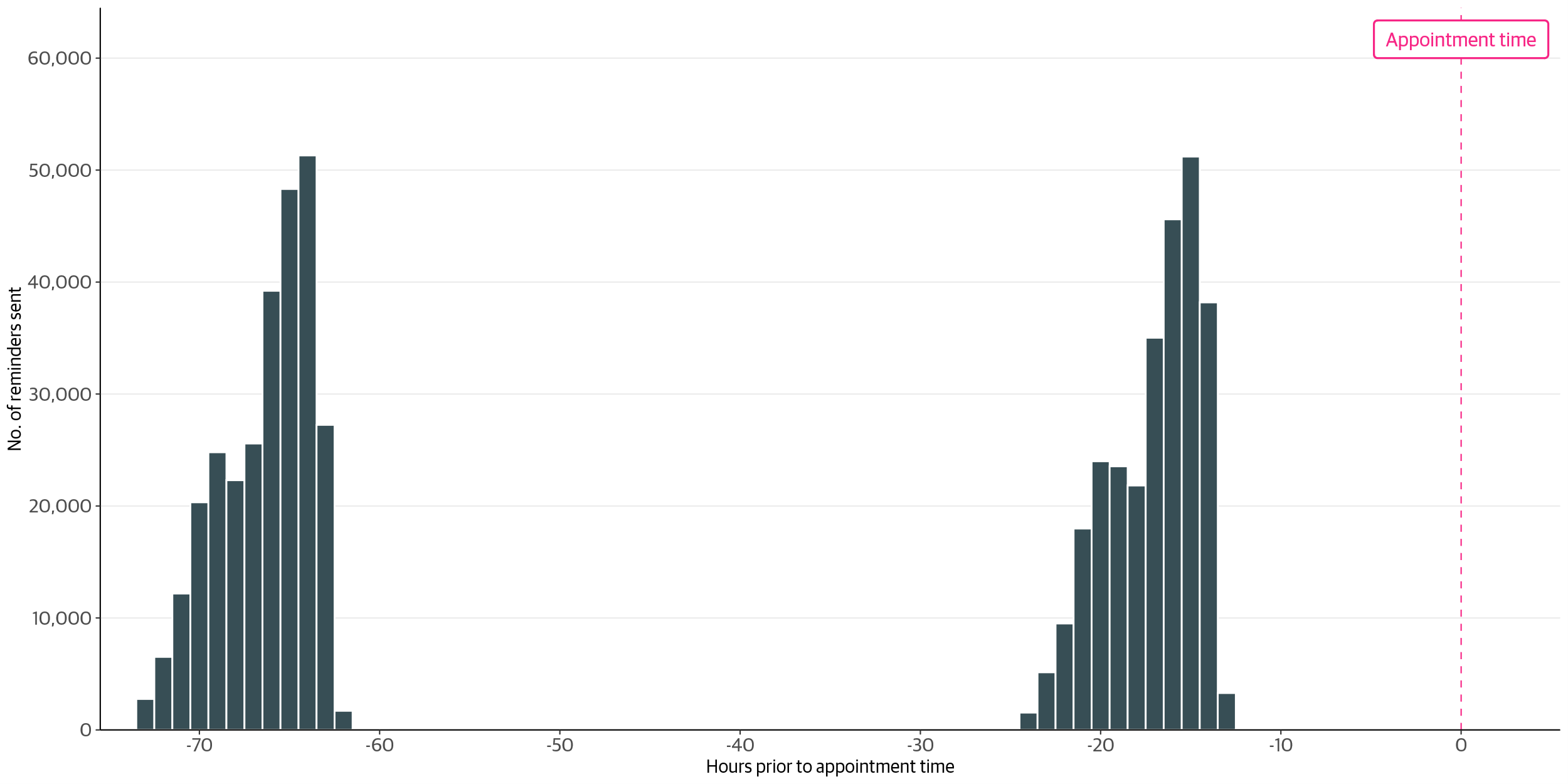

**Figure 1 Suppl Material. Number of messages sent by hours prior to appointment.**

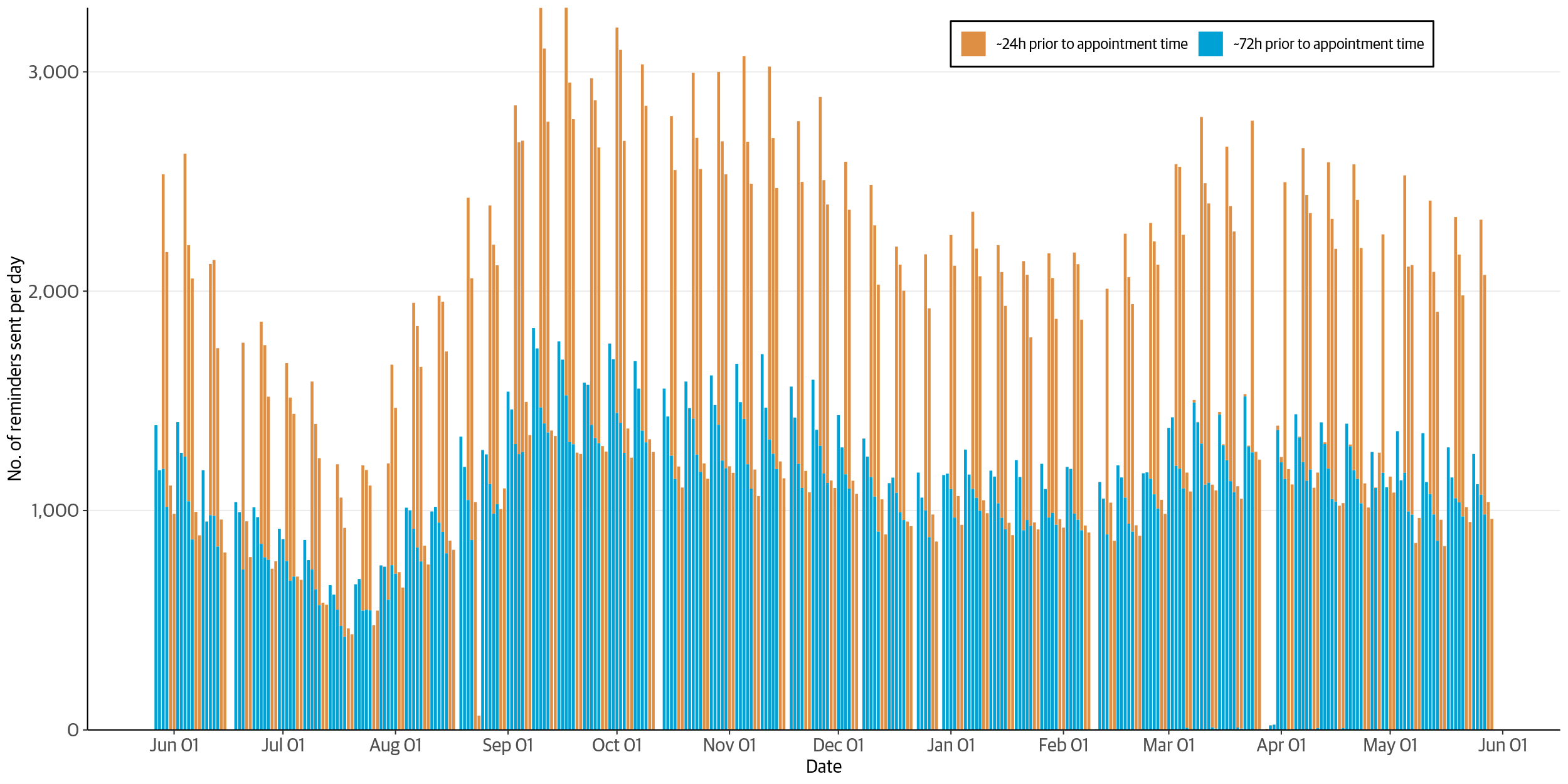

**Figure 2 Suppl Material. Distribution of reminders sent during the study period**

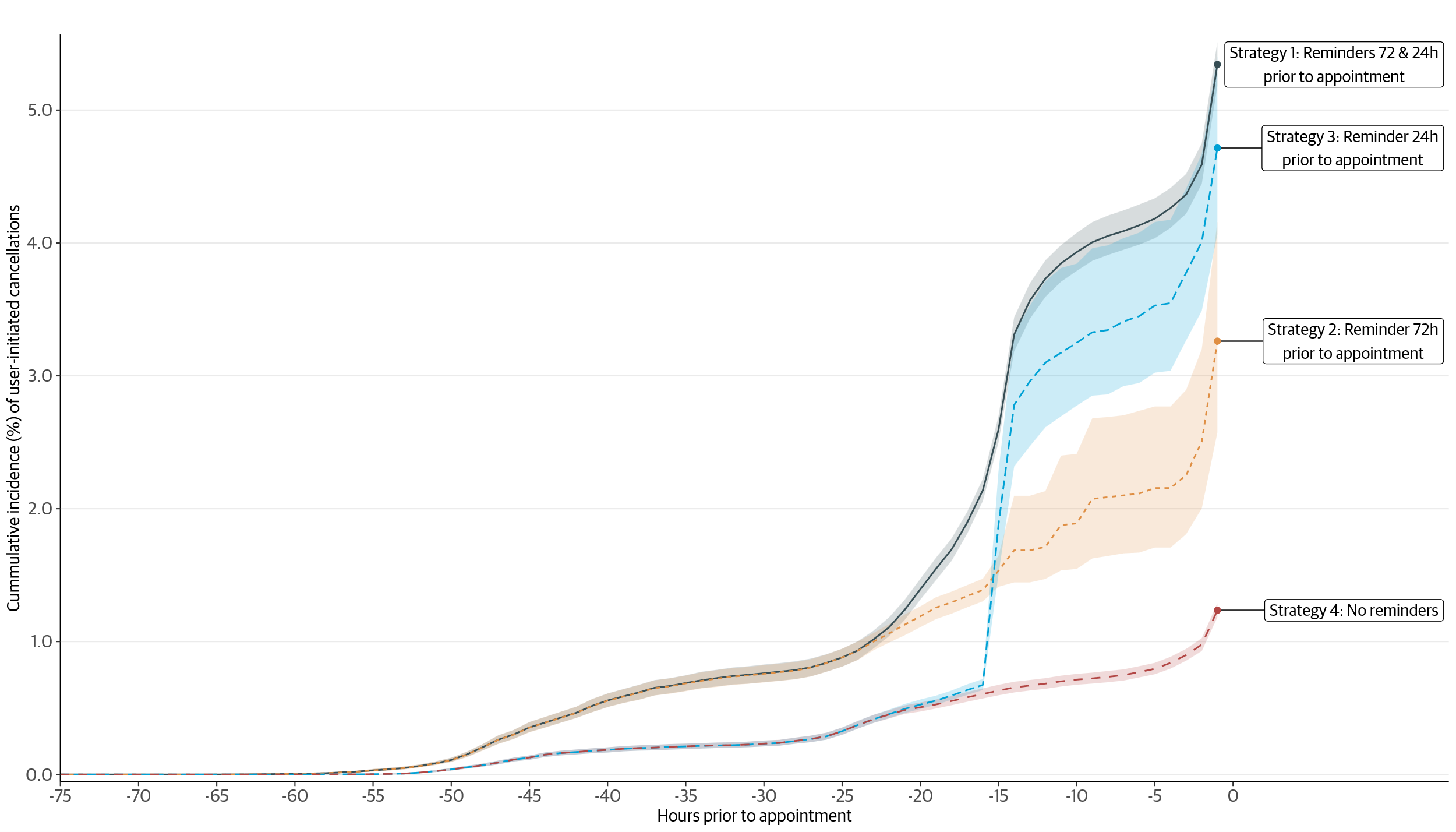

**Figure 3 Suppl Material. Estimated 75-hours risk of user-initiated cancellation by treatment group, Buenos Aires Public Health System data (June 2023-May 2024). Shaded areas represent pointwise 95% confidence intervals.**

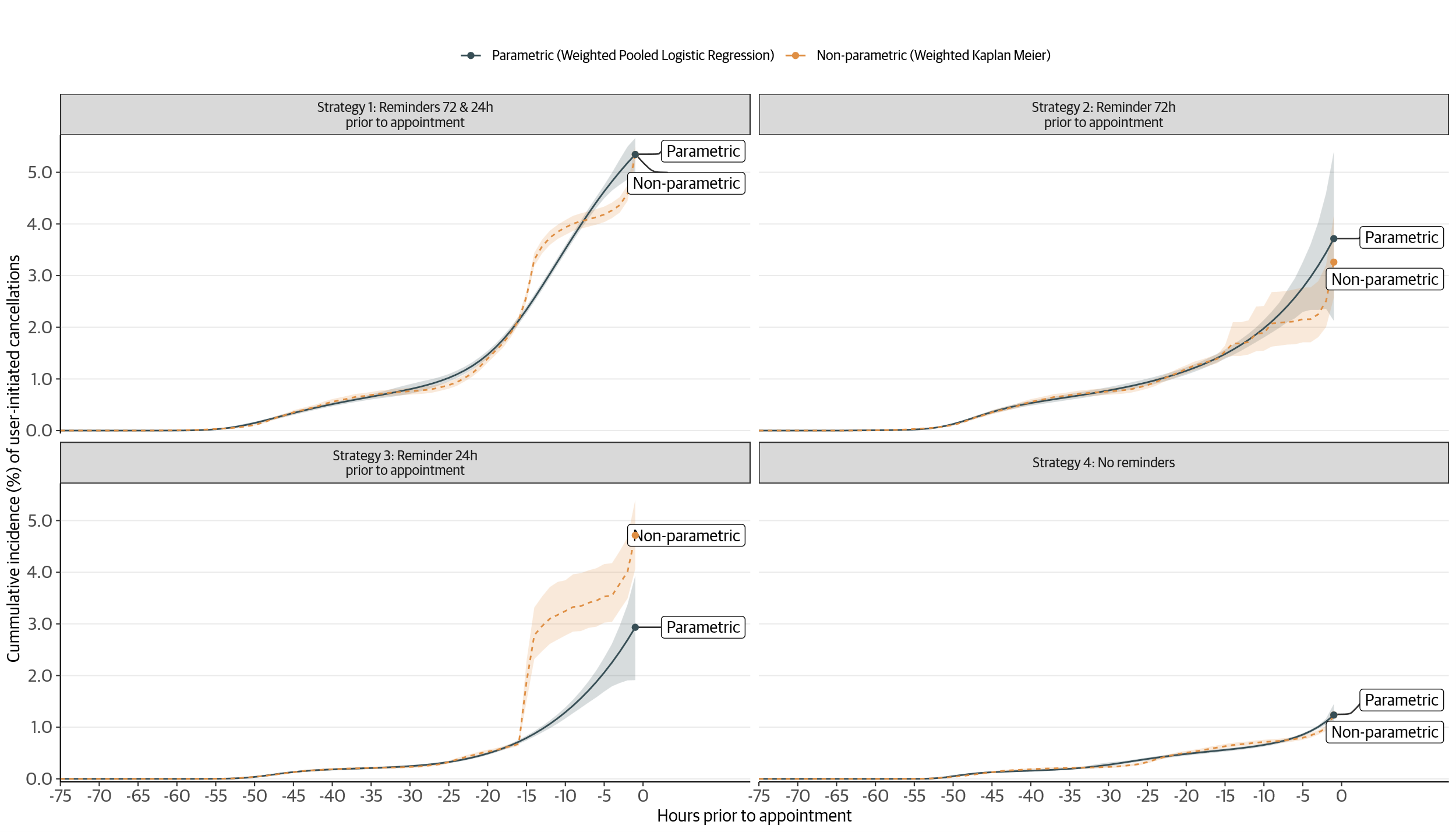

**Figure 4 Suppl Material. Sensitivity analysis:** Parametric vs non-parametric cumulative incidence estimation.

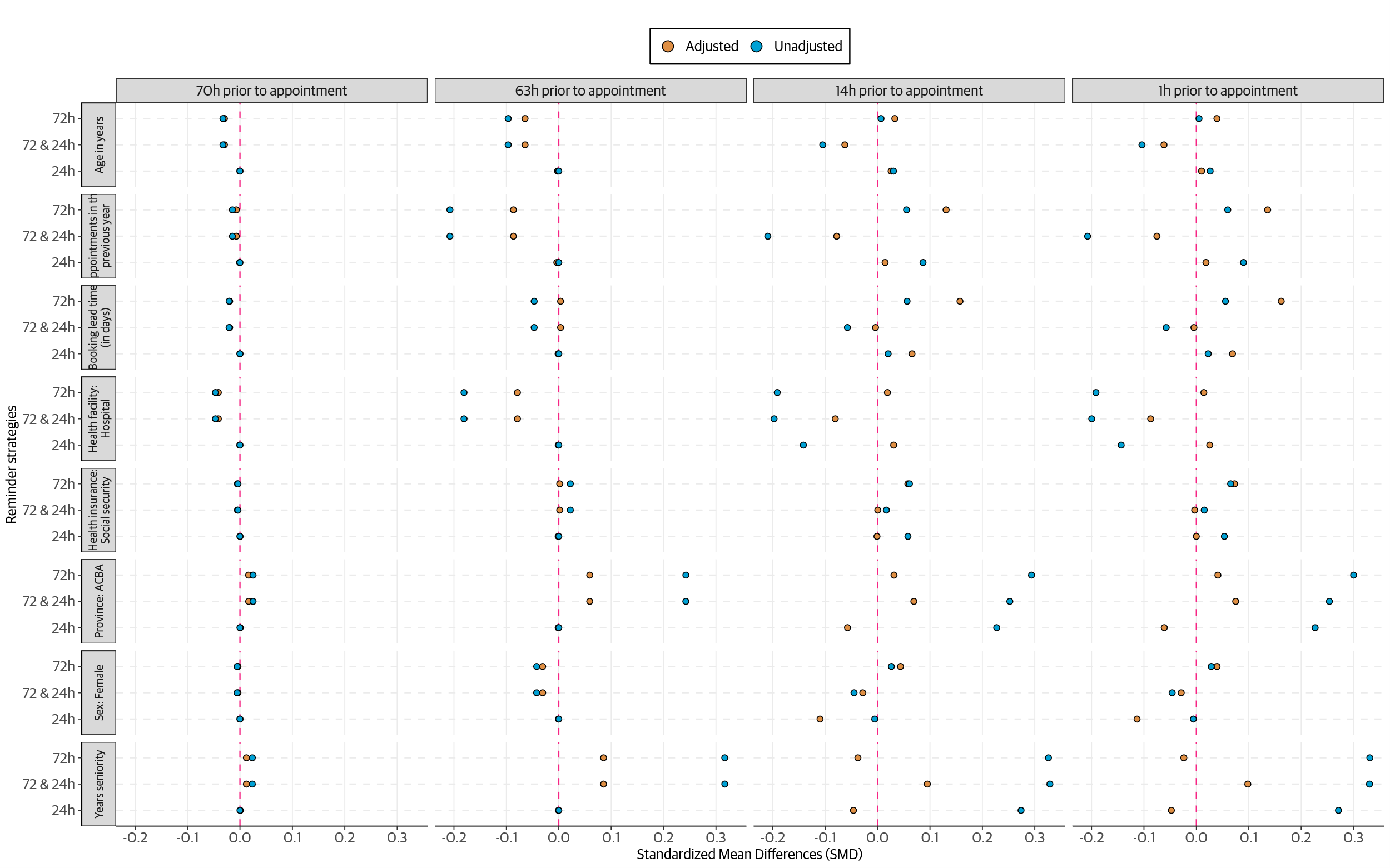

**Figure 5 Suppl Material. Standardized Mean Differences per reminder strategy compared to control arm.**

1. Turnos en hospitales y establecimientos de salud. [cited 7 Feb 2024]. Available: https://buenosaires.gob.ar/salud/hospitales-y-establecimientos-de-salud/turnos-en-hospitales-y-establecimientos-de-salud
2. Buenos Aires Ciudad - Gobierno de la Ciudad Autónoma de Buenos Aires. [cited 20 Dec 2025]. Available: https://buenosaires.gob.ar/innovacionytransformaciondigital/boti
